# Predictors and risk factors of short-term and long-term outcomes among women with gestational diabetes mellitus (GDM) and their offspring: Moving toward precision prognosis?

**DOI:** 10.1101/2023.04.14.23288199

**Authors:** Zhila Semnani-Azad, Romy Gaillard, Alice E Hughes, Kristen E. Boyle, Deirdre K. Tobias, Wei Perng, ADA/EASD PMDI

## Abstract

As part of the American Diabetes Association Precision Medicine in Diabetes Initiative (PMDI) – a partnership with the European Association for the Study of Diabetes (EASD) – this systematic review is part of a comprehensive evidence evaluation in support of the 2^nd^ International Consensus Report on Precision Diabetes Medicine. Here, we sought to synthesize evidence from empirical research papers published through September 1^st^, 2021 to evaluate and identify prognostic conditions, risk factors, and biomarkers among women and children affected by gestational diabetes mellitus (GDM), focusing on clinical endpoints of cardiovascular disease (CVD) and type 2 diabetes (T2D) among women with a history of GDM; and adiposity and cardiometabolic profile among offspring exposed to GDM *in utero.* We identified a total of 107 observational studies and 12 randomized controlled trials testing the effect of pharmaceutical and/or lifestyle interventions. Broadly, current literature indicates that greater GDM severity, higher maternal body mass index, belonging to racial/ethnic minority group; and unhealthy lifestyle behaviors would predict a woman’s risk of incident T2D and CVD, and an unfavorable cardiometabolic profile among offspring. However, the level of evidence is low (Level 4 according to the Diabetes Canada 2018 Clinical Practice Guidelines for diabetes prognosis) largely because most studies leveraged retrospective data from large registries that are vulnerable to residual confounding and reverse causation bias; and prospective cohort studies that may suffer selection and attrition bias. Moreover, for the offspring outcomes, we identified a relatively small body of literature on prognostic factors indicative of future adiposity and cardiometabolic risk. Future high-quality prospective cohort studies in diverse populations with granular data collection on prognostic factors, clinical and subclinical outcomes, high fidelity of follow-up, and appropriate analytical approaches to deal with structural biases are warranted.

## 1. INTRODUCTION

Gestational diabetes mellitus (GDM) is the most common metabolic disorder of pregnancy, affecting 6-12% of pregnancies globally ^1, 2^. A diagnosis of GDM indicates a state of maternal hyperglycemia due to insufficient insulin secretion and/or insulin resistance that is associated not only with risk of acute pregnancy complications, but also carries implications for the long-term health of the mother and offspring. For instance, in women, GDM is associated with up to 70% higher cumulative incidence of type 2 diabetes (T2D) following the index pregnancy ^3^ and two-fold greater risk of cardiovascular disease (CVD) than gravidas without GDM ^4^. Additionally, offspring exposed to GDM *in utero* have higher adiposity and a worse metabolic profile than their unexposed counterparts ^5^. The wide-ranging and intergenerational sequelae of GDM-affected pregnancies emphasize the importance of characterizing not only the short- and long-term consequences of this common pregnancy complication, but also, early bellwethers of such consequences to facilitate preventive intervention of such comorbidities and complications – a concept known as disease *prognosis*.

Recent technological advancements have improved the capacity to comprehensively assess physiology. In turn, these developments facilitated the ability to harness metabolic heterogeneity – the phenomenon of interest to precision medicine ^6^, by which similar exposures and risk factors yield differential health sequelae across individuals. In the context of GDM prognosis, this effort requires the identification of prognostic conditions and biomarkers among women with a history of GDM and/or their children who were exposed to GDM *in utero* that may serve as both causal and non-causal indicators of future health risks.

Recognizing the relevance of metabolic heterogeneity in accurate and precise assessment of metabolic disease prediction, diagnosis, treatment, and prognosis, the *Precision Medicine in Diabetes Initiative* (PMDI) was established in 2018 by the American Diabetes Association (ADA) in partnership with the European Association for the Study of Diabetes (EASD). The ADA/EASD PMDI includes global thought leaders in precision diabetes medicine who are working to address the burgeoning need for better diabetes prevention and care through precision medicine ^7^. This Systematic Review is written on behalf of the ADA/EASD PMDI as part of a comprehensive evidence evaluation in support of the 2^nd^ International Consensus Report on Precision Diabetes Medicine ^1^.

Here, we synthesize evidence from empirical research papers published through September 1^st^, 2021 to evaluate and identify prognostic conditions, risk factors, and biomarkers among women and children affected by GDM pregnancies, focusing on clinical endpoints of CVD and T2D among women with a history of GDM; and adiposity and cardiometabolic profile among offspring exposed to GDM *in utero*.

## 2. METHODS

### 2.1. Systematic review protocol development

We developed a systematic review protocol to comprehensively include and evaluate individual research studies reporting on risk factors for long-term clinical outcomes in women with GDM and a range of cardiometabolic health and anthropometric outcomes in GDM-exposed offspring. Our strategy aimed to identify two broad categories of empirical studies: (1) populations of women with a history of prior GDM that investigated additional exposures or risk factors for incident postpartum T2D or CVD; (2) populations comprising offspring exposed to GDM *in utero*. Studies including pregnancies unaffected by GDM were eligible only if results among GDM subgroups were reported.

Prognostic factors of interest – hereafter also referred to as “exposures” – included sociodemographic and environmental factors, lifestyle and behavioral characteristics, traditional clinical traits, and novel ‘omics biomarkers. We considered these prognostic factors during the perinatal/postpartum periods and across the lifecourse for both the mothers and offspring. Maternal outcomes of interest were incident T2D or CVD, including study-specific composites of clinical cardiovascular events, non-fatal and fatal myocardial infarction or stroke, and chronic kidney disease (CKD). For offspring, we were interested in outcomes reported 12 weeks of age and older, and limited to anthropometrics, glycemic and cardiometabolic traits or biomarkers, and incident metabolic syndrome (MetS), T2D, or CVD.

### 2.2. Data sources, search strategy, and screening criteria

We published our search strategy and systematic review protocol to PROSPERO CRD42021276094 ^8^. We developed search terms for Medline EMBASE, and Cochrane CENTRAL (**Supplemental Table 1**) for eligible citations published in English language from January 1^st^, 1990 through September 30^th^, 2021. References of accepted manuscripts and relevant systematic reviews published within the past 2 years were screened to identify additional citations. We included prospective and retrospective observational studies identifying factors with incident outcomes of interest in women or offspring exposed to GDM. We excluded cross-sectional analyses among populations with prevalent disease outcomes or traits. While studies could include non-GDM exposed pregnancies, those without subgroup findings exclusively among GDM pregnancies were excluded. We also included interventions prospectively comparing effects of a treatment assignment on the outcome. Exclusion criteria comprised studies with outcomes <12 weeks postpartum, maternal studies reporting only intermediate phenotypes, glycemic traits, or cardiometabolic biomarkers, and studies in offspring that only assessed endpoints outside of the cardiometabolic outcomes of interest (e.g., neurodevelopment, allergic disease). Using these, two independent reviewers conducted screening at the title abstract level. For accepted citations, two independent reviewers implemented screening of the full manuscripts. Conflicts at all screening stages were resolved by a third reviewer. All screening was conducted in the Covidence online systematic review tracking platform.

### 2.3. Data extraction and synthesis of results

We developed and piloted a data extraction template for eligible manuscripts. Data included manuscript information, study level details and design, population enrollment and characteristics, exposure and outcome ascertainment and diagnosis criteria, follow-up time of outcome assessment since index GDM pregnancy and other pertinent details. We indicated the population in which outcomes were assessed (e.g., maternal, offspring, or both), and recorded the exposures that were investigated in four broad categories: (i) social/environmental/genetics factors across the life course; (ii) all factors in perinatal/postpartum window; (iii) long-term maternal exposures; and (iv) long-term offspring exposures.

### 2.4. Quality assessment (risk of bias) and synthesis

We assessed the quality of each study using the Joanna Briggs Institute’s (JBI) critical appraisal tools for cohort studies and randomized controlled trials (RCTs) ^9^. For cohort studies, we assessed quality based on 11 items which evaluated population recruitment, exposure and outcome ascertainment, confounding, statistical methodology, and follow-up. For the RCTs, the JBI criteria evaluated 13 items which assessed selection and allocation, intervention, administration, outcome ascertainment, follow-up, and statistical analysis. Each JBI item was categorized as ‘Yes’, ‘No’, ‘Unclear’ or ‘Not applicable’ following the described guidelines. Any uncertainty in assessment was further discussed by the full research team.

### 2.5. Overall evidence certainty assessment and synthesis

The certainty of evidence was determined using the Diabetes Canada 2018 Clinical Practice Guidelines for studies ^10^. Levels were based on study design and criteria focused on inception cohort of patients presenting GDM but without outcomes of interest, inclusion/exclusion reproducibility, follow-up of at least 80% of participants, adjustment for confounding factors, and reproducible outcome measures. Scoring ranged from level 1 to 4, with Level 1 indicating the highest certainty of evidence and Level 4 indicating the lowest certainty of evidence. Details on the criteria and guidelines are presented in **Supplemental Table 2**.

## 3. RESULTS

**Figure 1** displays results of the systematic search and article inclusion process. Of the 8,141 studies identified, five were excluded due to duplication. Another 7,770 were excluded following title and abstract review. The remaining 341 studies were reviewed in full. A total of 119 studies met the inclusion criteria for this review.

**FIGURE 1.**
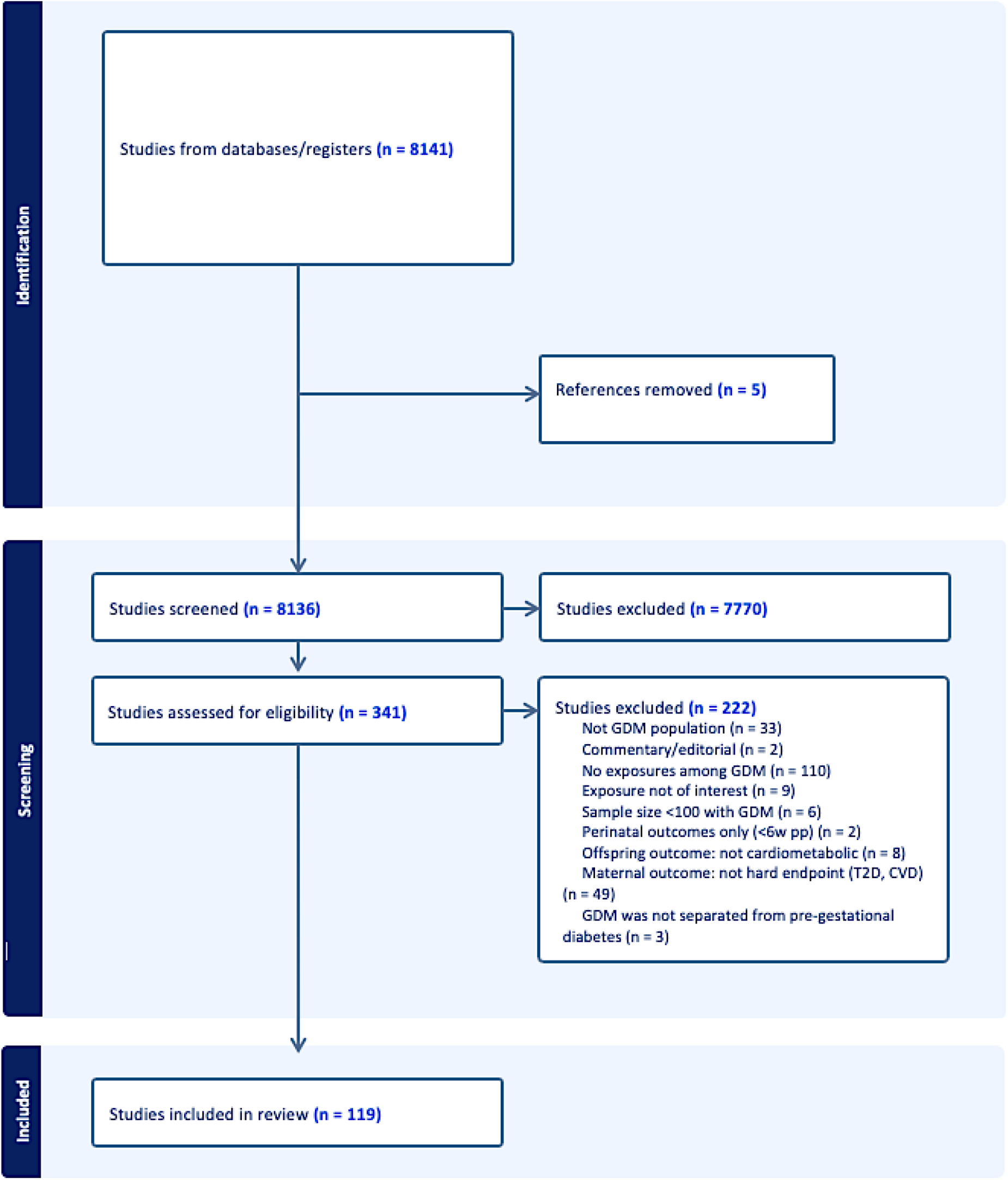
Systematic review screening and attrition diagram.

**Table 1** outlines the summary of studies included in this systematic review. Of the 119 studies identified, 107 were observational studies and 12 were RCTs. Of the observational studies, 57 studies ^2–55^ reported on maternal outcomes and 51 studies ^56–106^ reported on offspring outcomes. Of the RCTs, three evaluated maternal outcomes ^107–109^ and nine assessed offspring outcomes ^61, 91, 110–116^. Studies included data from primarily from Caucasian ethnicities with publications mainly from North American (n = 25) and European (n = 39) countries. The sample sizes of the eligible studies ranged from 26 to 23 302.

**TABLE 1.**
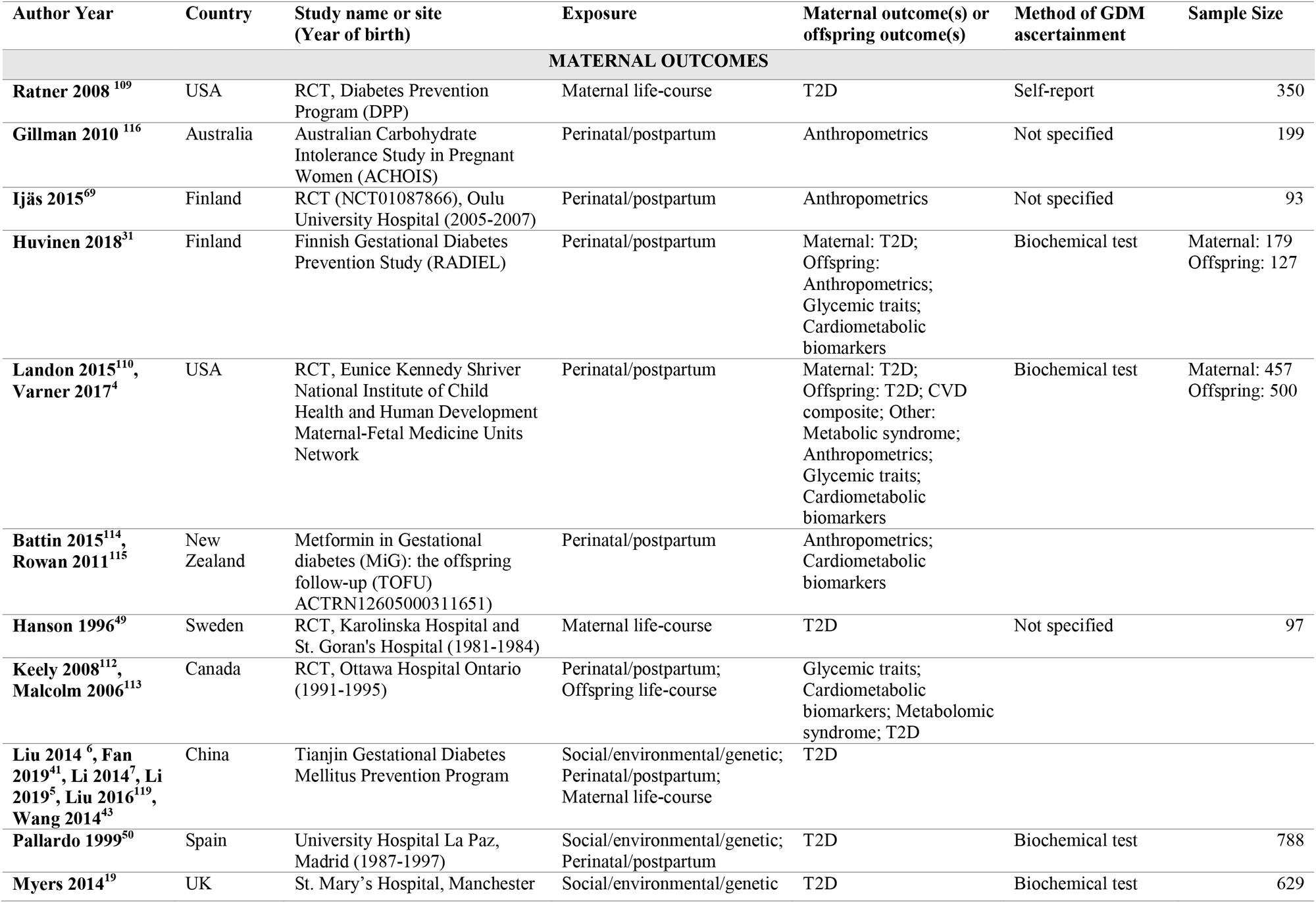
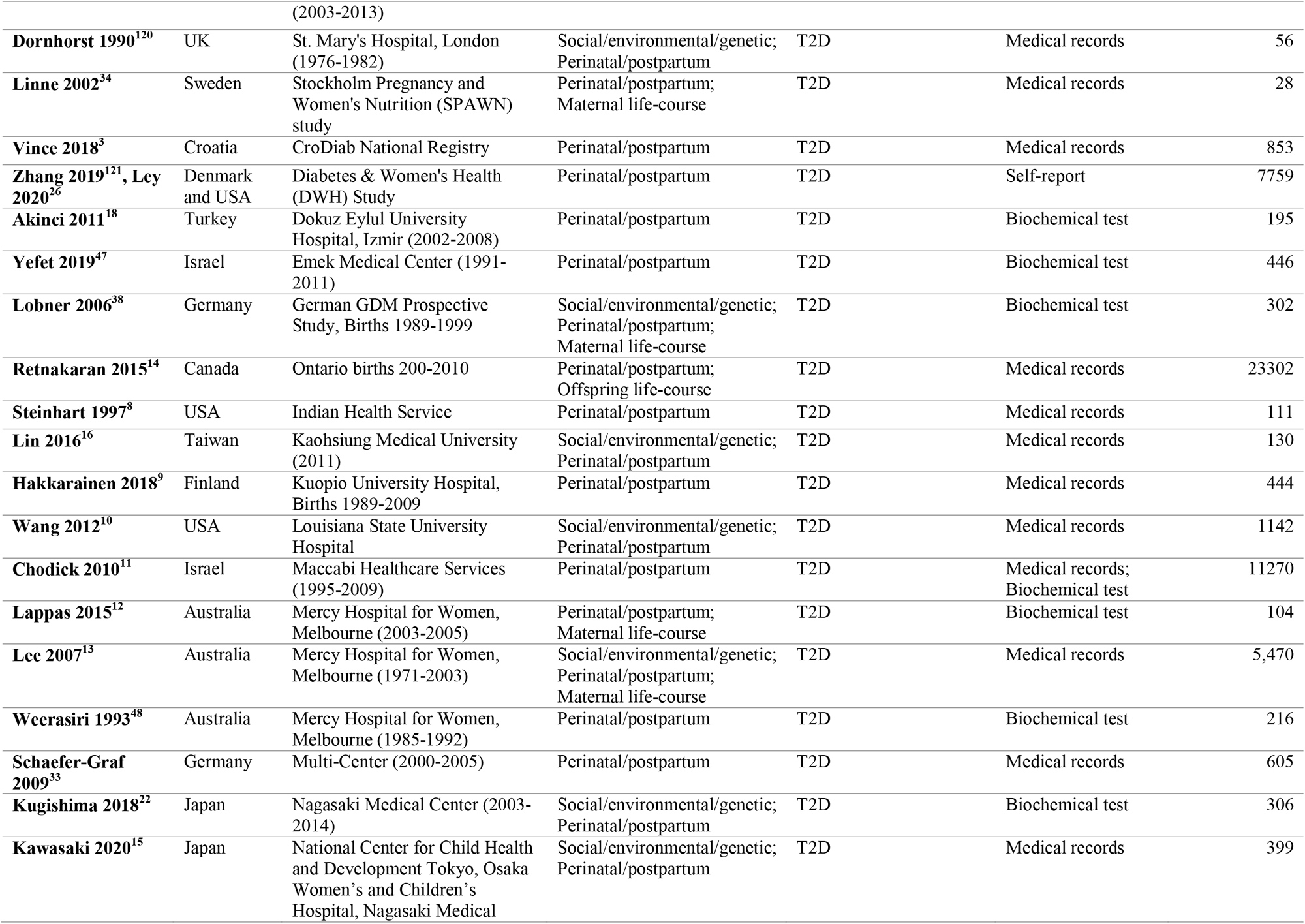
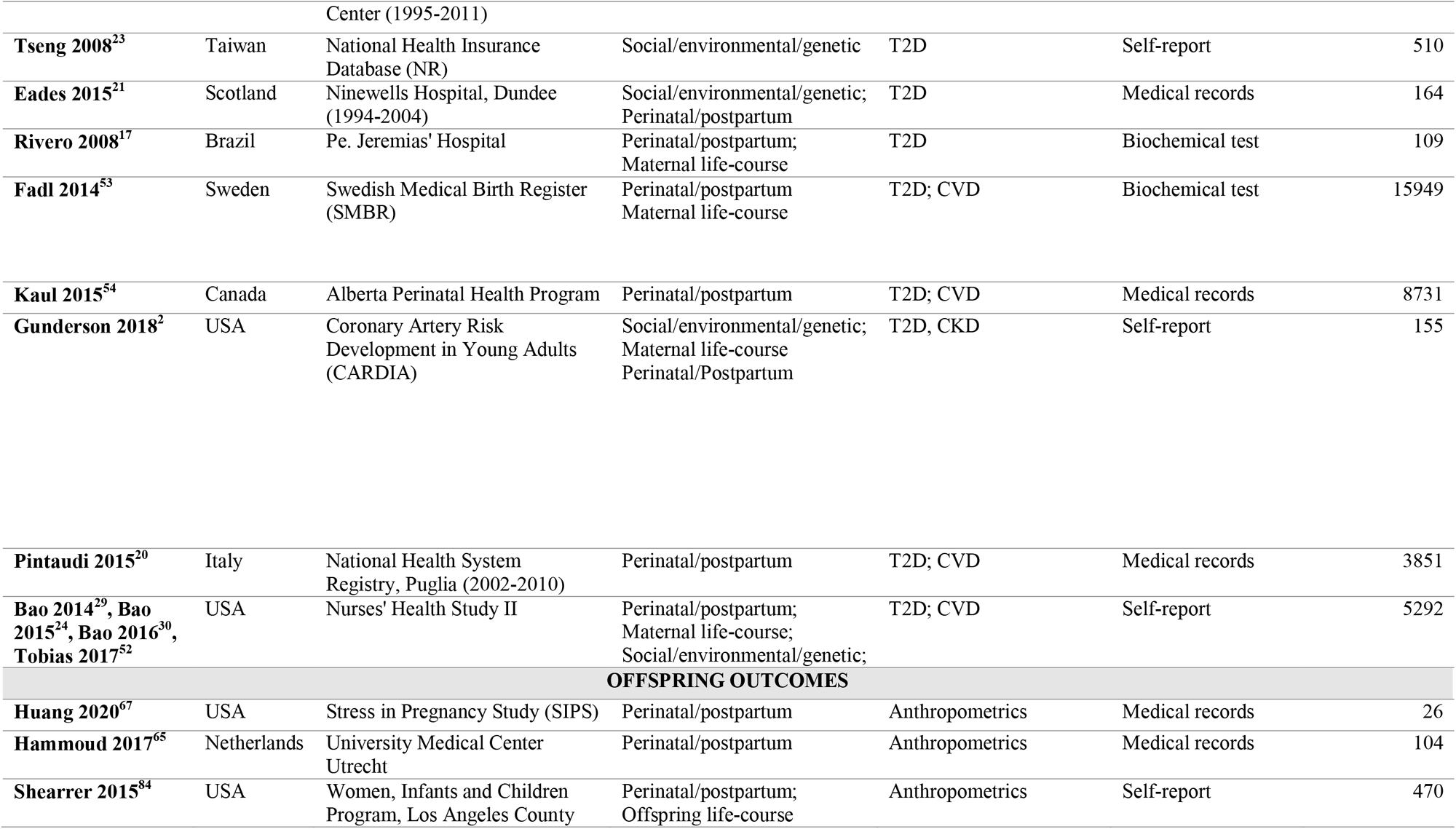
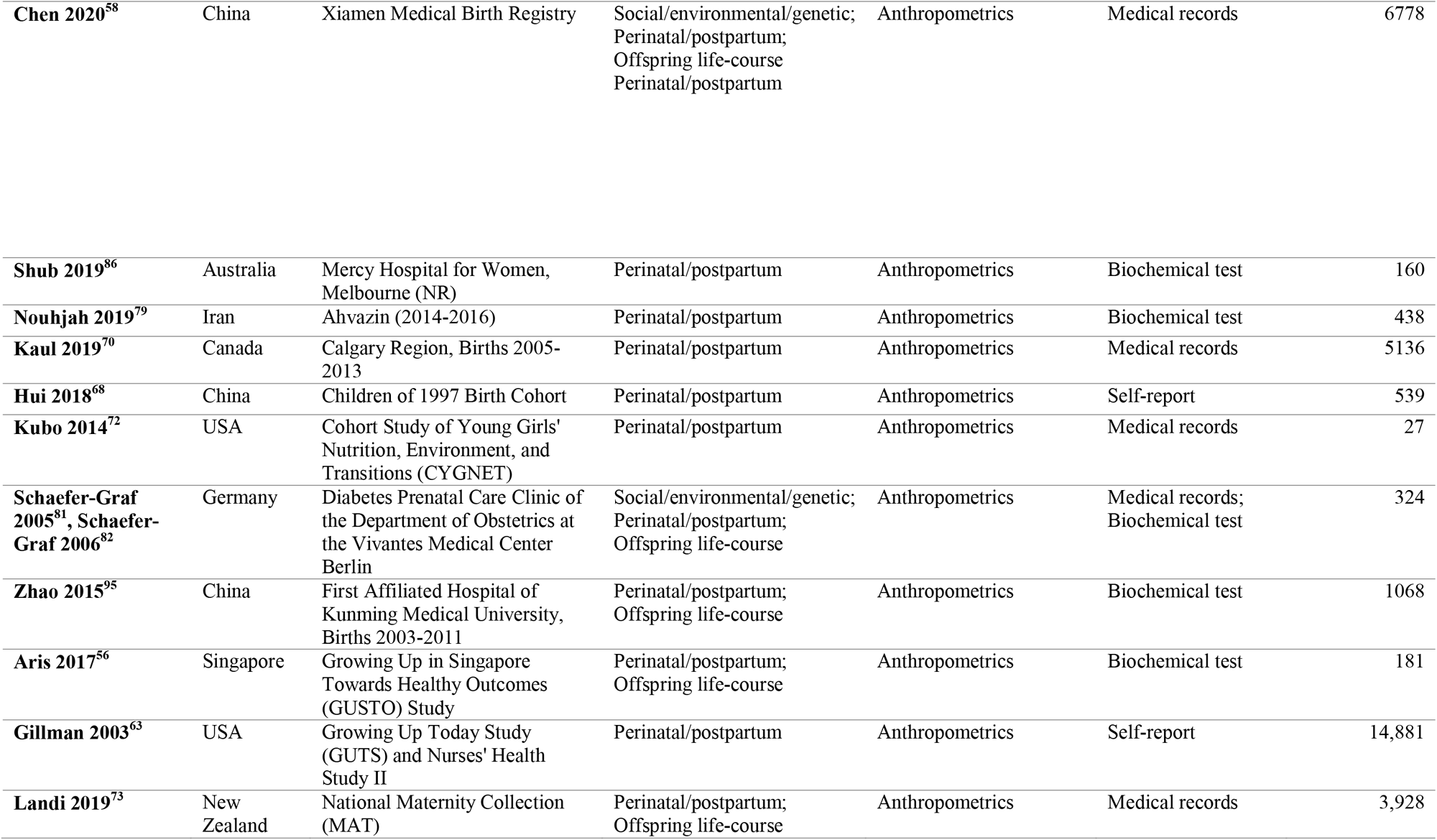
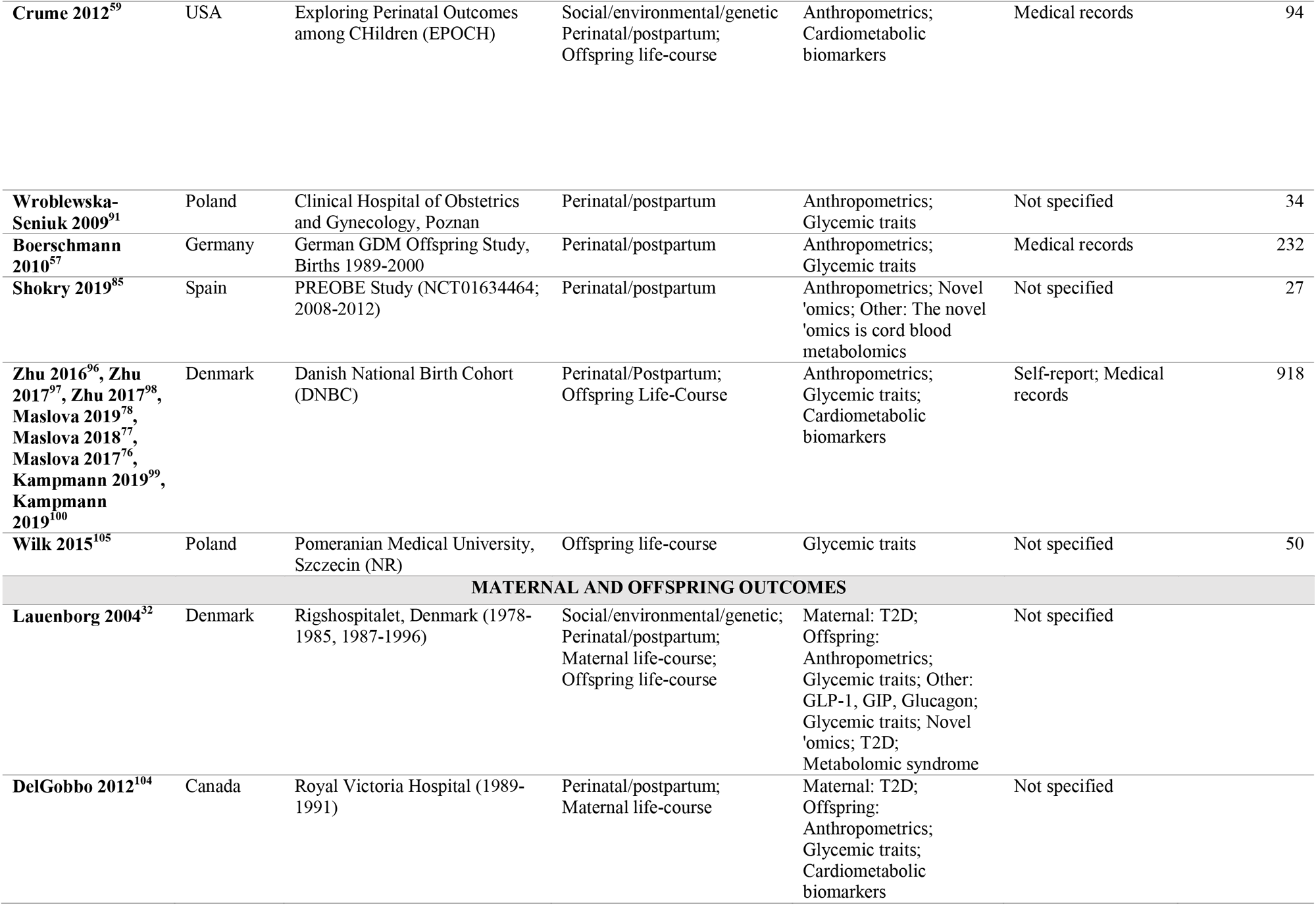
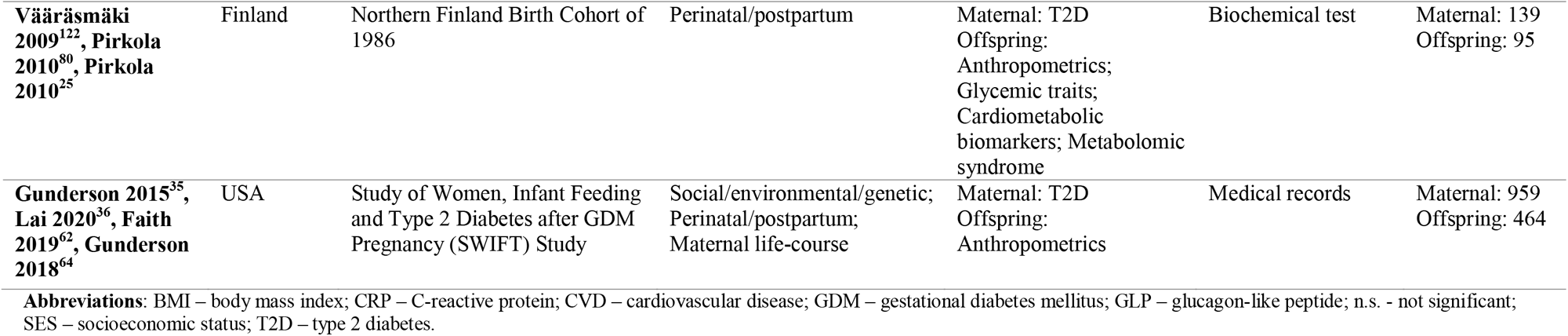
Summary of studies included in this systematic review, stratified by maternal, offspring, and maternal and offspring outcomes.

### 3.1. Maternal outcomes

#### 3.1.1. Maternal type 2 diabetes

Fifty-one observational studies ^11–61^ (**Table 2**) and three RCTs ^62–64^ (**Table 3**) assessed socio-demographic, lifestyle, clinical and pregnancy characteristics associated with the risk of T2D among GDM women. The most commonly-studied characteristics were maternal BMI and GDM severity. Twenty-four observational studies showed that a higher BMI, measured prior to pregnancy, pregnancy or during the lifecourse was associated with a higher risk of T2D. Two observational studies further showed that a higher weight gain in the postpartum period also increased the risk of T2D. Fifteen studies assessed the role of GDM severity on the risk of developing T2D and showed that more severe GDM, measured by either clinical markers or need for insulin, was associated with a higher risk of developing T2D (**Table 2**). One study was a RCT; all other studies were observational. Fewer studies explored the role of lifestyle/behavioral, pregnancy, and clinical characteristics. Six studies explored the role of race/ethnicity in risk of T2D, two of which showed no significant associations (**Table 2**). Four studies suggested that the risk was higher among women with non-European ancestry. With regards to pregnancy characteristics, findings were inconsistent with three of six observational studies reporting an association of a complicated pregnancy with higher risk of T2D. The pregnancy complications assessed varied across reports. Six studies explored the role of parity, of which three showed that higher parity was associated with a higher risk of T2D. Four observational studies showed that breastfeeding was associated with reduced risk of developing T2D in later life. Three studies including one RCT focused on various healthy dietary patterns and the risk of T2D among GDM women but showed inconsistent results. Seven small studies used novel clinical biomarkers, including metabolomics, lipidomics, and diabetes auto-antibodies, and found positive associations with T2D.

**TABLE 2.**
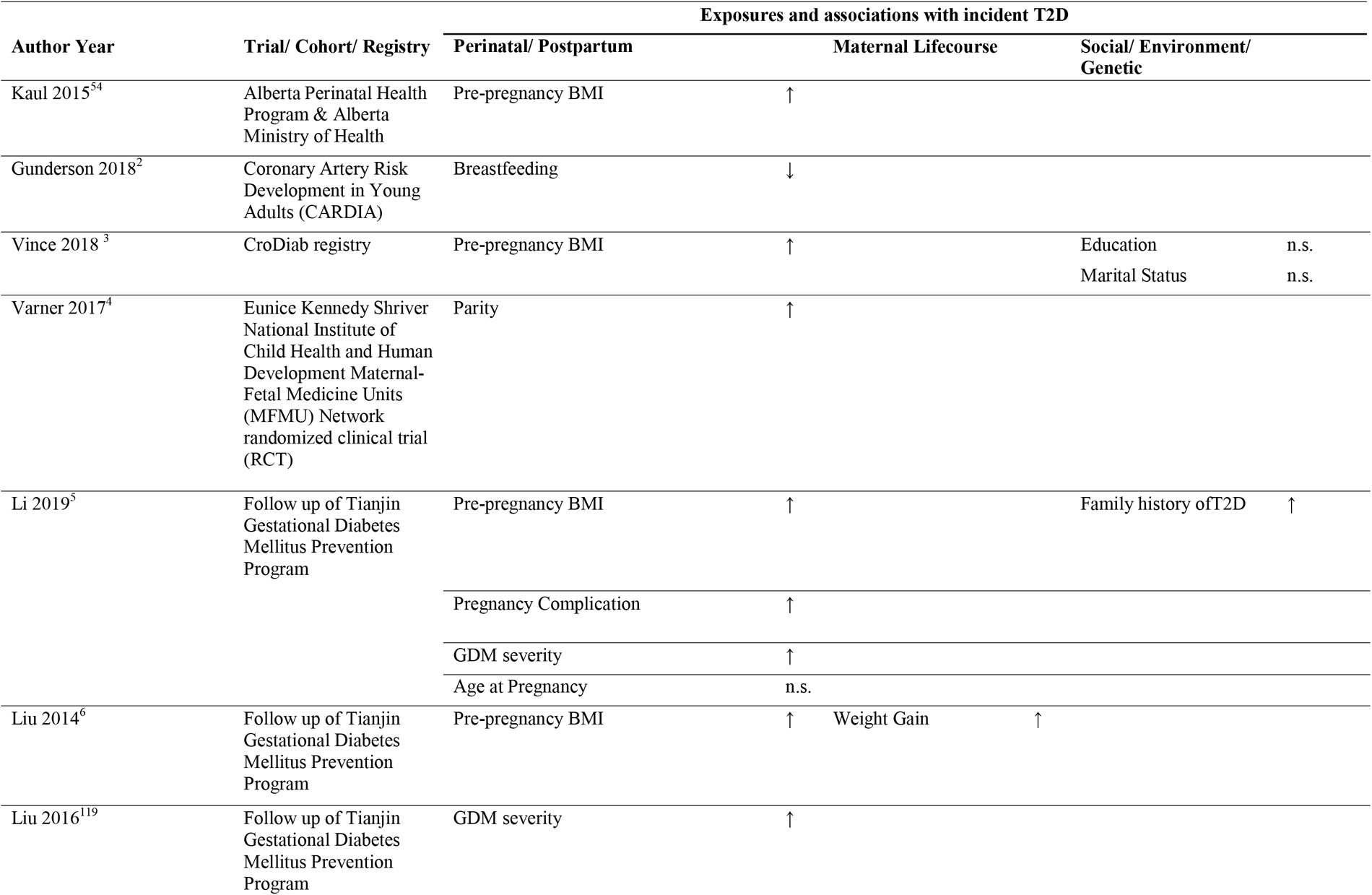
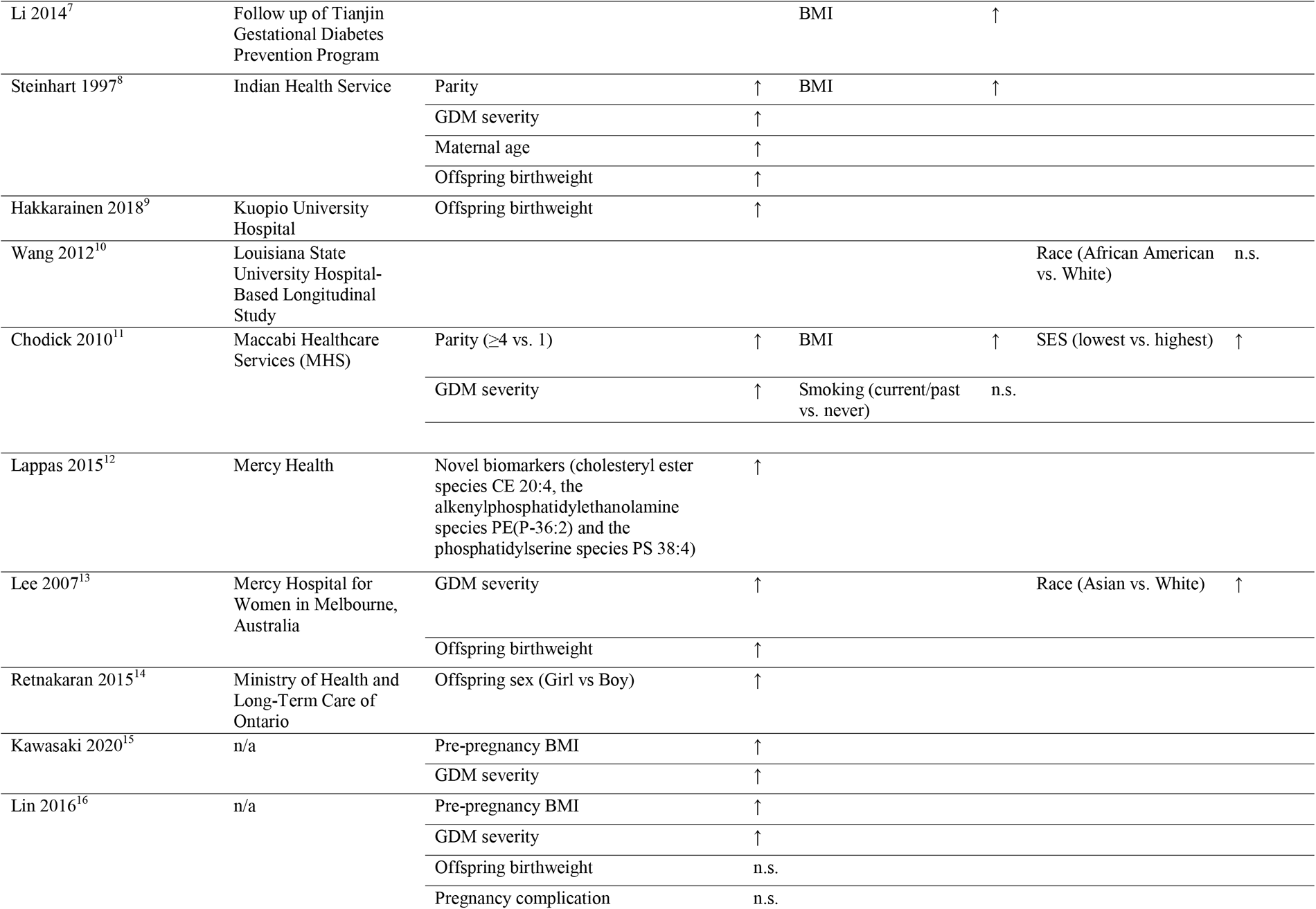
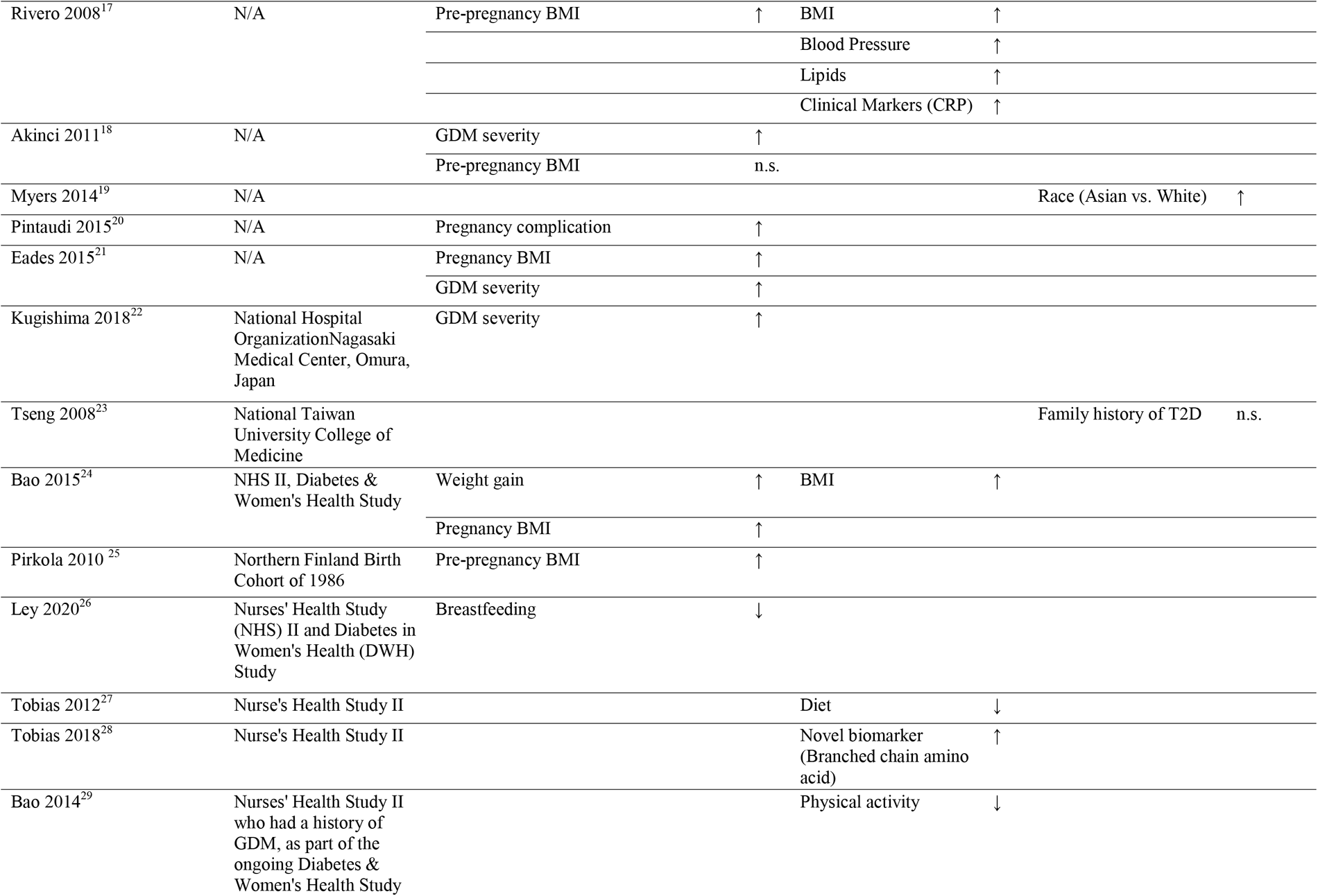
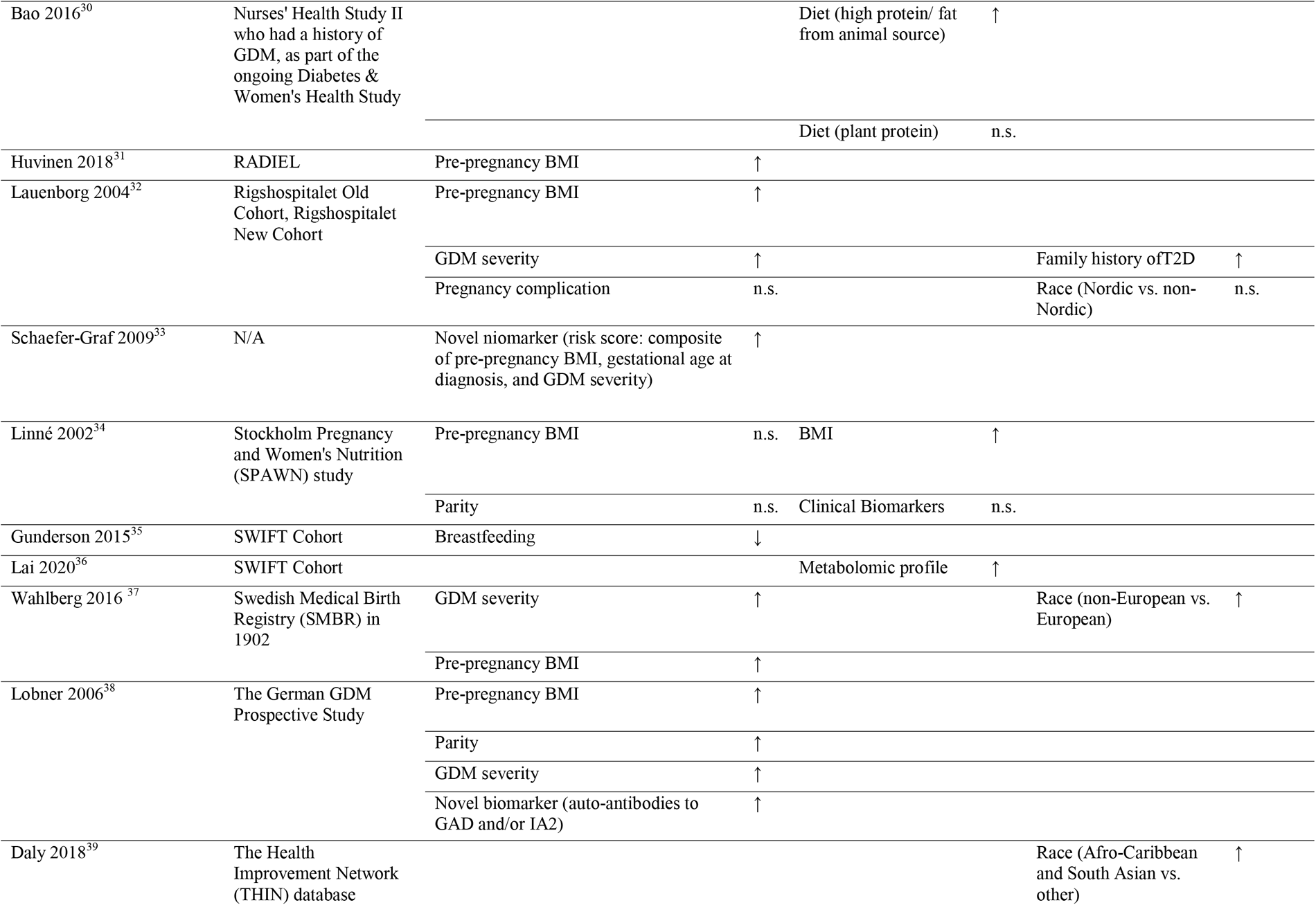
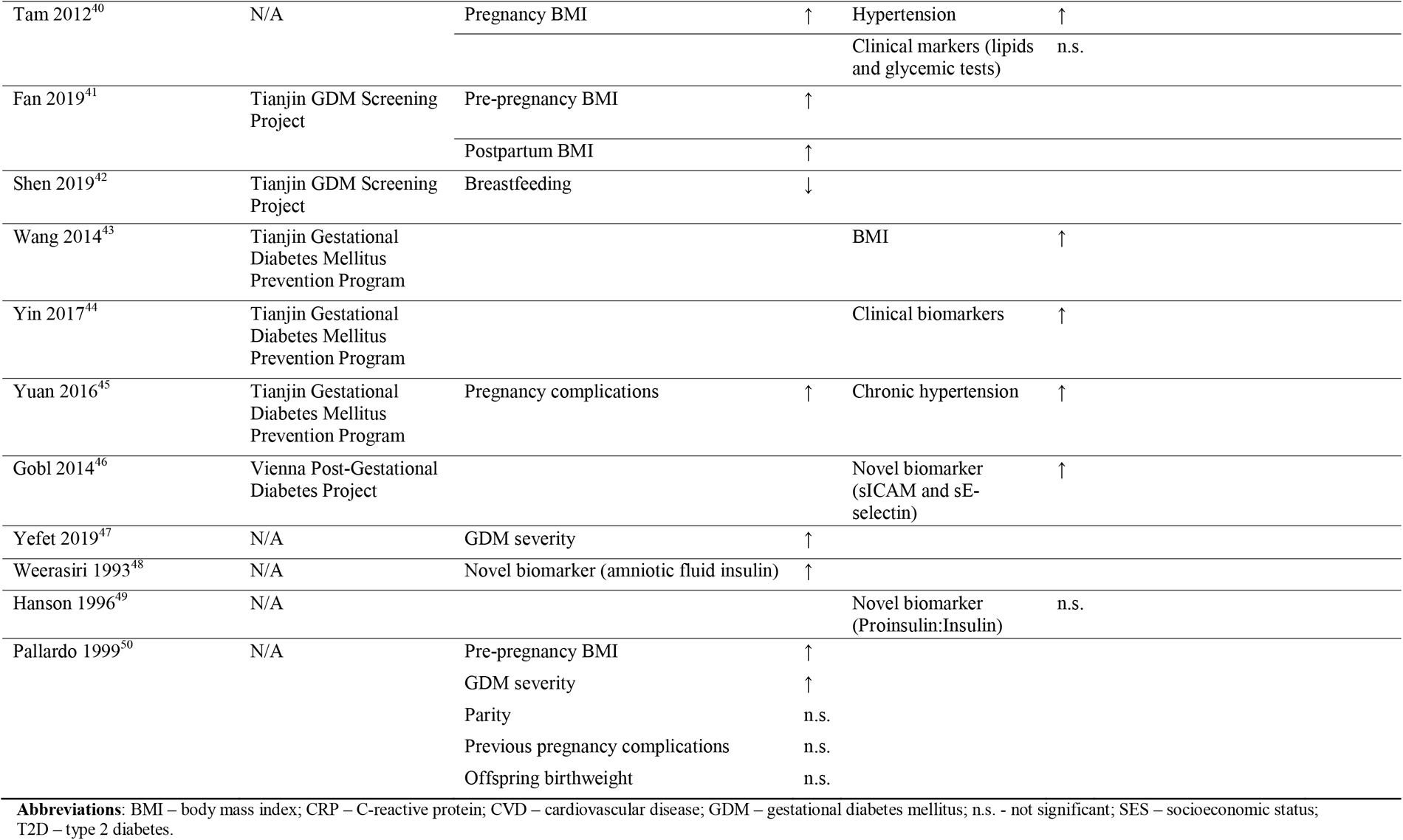
Observational studies evaluating prognostic factors associated with incident T2D among women with a history of GDM (n=51).

**TABLE 3.**
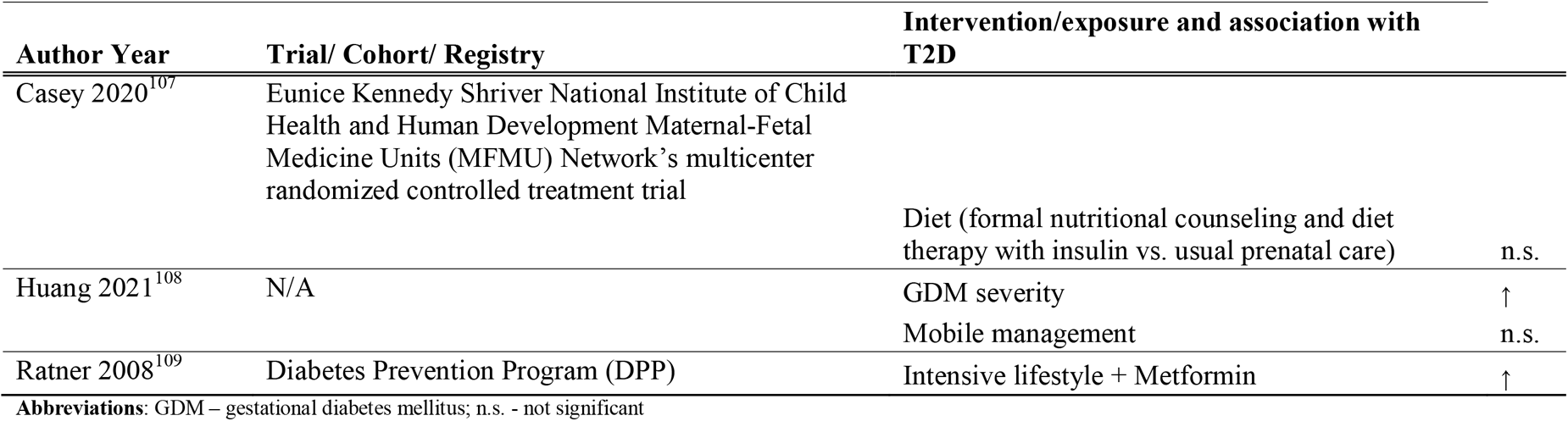
Randomized controlled trials (RCT) evaluating prognostic factors associated with incident type 2 diabetes (T2D) among women with a history of GDM (*n*=3).

#### 3.1.2. Maternal cardiovascular diseases

Six observational studies ^11, 31, 65–68^ (**Table 4**) explored the role of sociodemographic, lifestyle, and pregnancy characteristics in future risk of CVD among women with GDM. Three studies ^11, 66, 67^ identified maternal BMI, before or during pregnancy, as a determinant of CVD risk, in which overweight and obese women with GDM have a higher risk of future CVD as compared to normal weight women with GDM. A study further showed that a healthy lifestyle – i.e., healthy diet, physical activity and non-smoking - was associated with a lower risk of CVD among women with GDM ^66^. Two studies showed that pregnancy complications, including gestational hypertension ^68^ and stillbirth ^31^, predicted risk of CVD among women of GDM. No significant effect modification was found by family history or chronic hypertension.

**TABLE 4.**
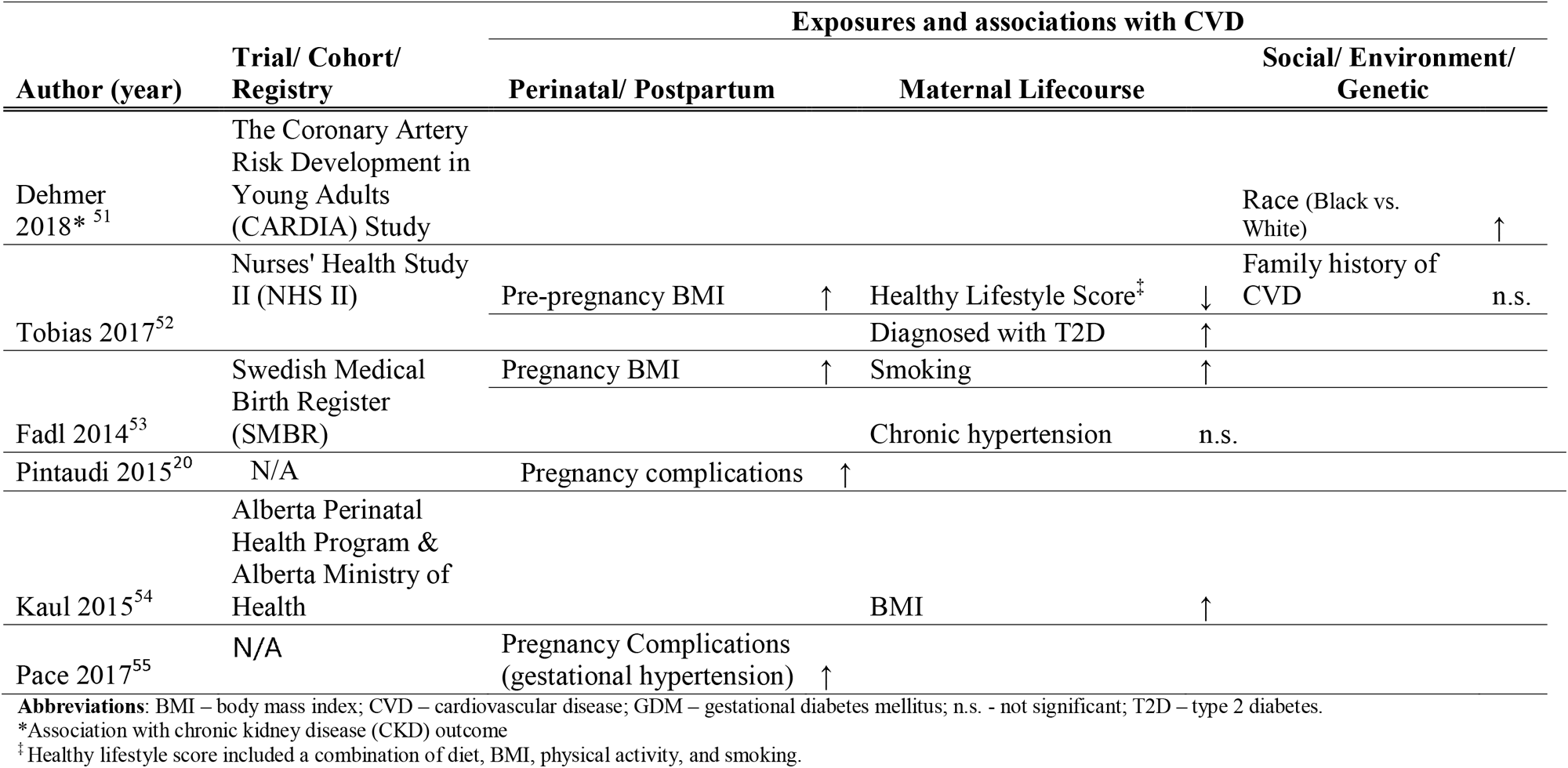
Observational studies evaluating prognostic factors associated with cardiovascular disease (CVD) among women with a history GDM (n=6).

#### 3.1.3. Quality of studies conducted and certainty of evidence in maternal populations

The quality of studies reviewed herein for prognostic factors indicative of risk of T2D or CVD is low and the overall certainty of evidence ranked between Levels 3 and 4 according to the Diabetes Canada 2018 Clinical Practice Guidelines ^10^. Most current literature were based on retrospective studies leveraging registry data and observational cohort studies, both of which are vulnerable to bias and an inability to attribute causation to prognostic factors due to residual confounding, reverse causation bias by pre-existing conditions, and other characteristics around time of pregnancy and GDM diagnoses.

### 3.2. Offspring outcomes

#### 3.2.1. Anthropometry and body composition

In comparison to the relatively large maternal literature, fewer studies have focused on prognostic factors associated with suboptimal offspring body composition among those exposed to GDM *in utero*. Forty-three observational studies ^56–98^ (**Table 5**) and four RCTs ^110, 113, 115, 116^ (**Table 6**) examined associations of sociodemographic, lifestyle, clinical and pregnancy characteristics associated with anthropometric outcomes offspring of GDM women. The most commonly-studied characteristics were maternal BMI, GDM severity, breastfeeding status, and birthweight, and were compared to anthropometric outcomes, specifically BMI and risk of overweight. Five observational studies showed that a higher maternal pre-pregnancy BMI was associated with an adverse body composition in the offspring, reflected by a higher BMI, waist circumference, or body fat mass, and a higher risk of overweight or obesity. Twelve studies assessed the associations of maternal GDM severity, measured by either clinical markers or need for insulin, with offspring body composition. Most of eight observational studies, only three indicated that more severe maternal GDM is associated with a higher offspring BMI and overweight risk. All four identified RCTs evaluated GDM severity and overall showed no significant association with offspring anthropometry or body composition. Four observational studies showed that a higher size at birth is associated with higher BMI and risk of overweight in GDM offspring, but four studies reported no significant associations. Two studies showed that offspring that a higher size at birth but with exposure to breastfeeding were associated with lower offspring BMI and reduced risk of overweight or obesity. With regards to breastfeeding status, only observational studies showed that exclusive breastfeeding and a longer duration of breastfeeding were associated with a lower offspring BMI and lower risk of overweight or obesity. Only four small studies used novel clinical biomarkers, including metabolomics in cord blood and gene expression in muscle and adipose tissue and showed variable associations with body composition. Furthermore, four independent studies using data from the Danish National Birth Cohort indicated that maternal diet consisting of fatty fish ^77^, refined grain ^98^, and sugar-sweetened beverage intake ^97^ were associated with higher offspring BMI, while protein intake did not show significant impact on offspring abdominal fat ^76^.

**TABLE 5.**
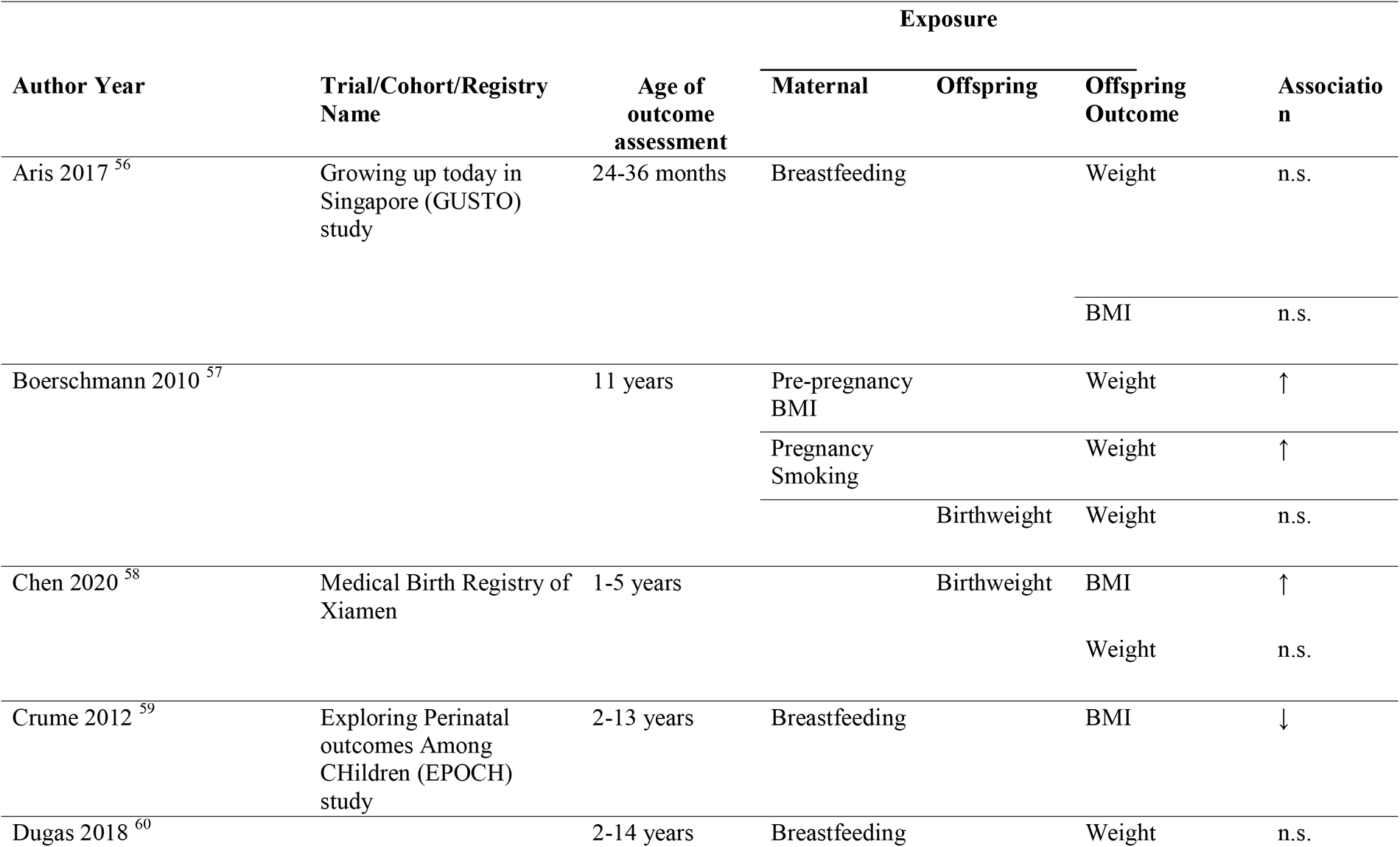
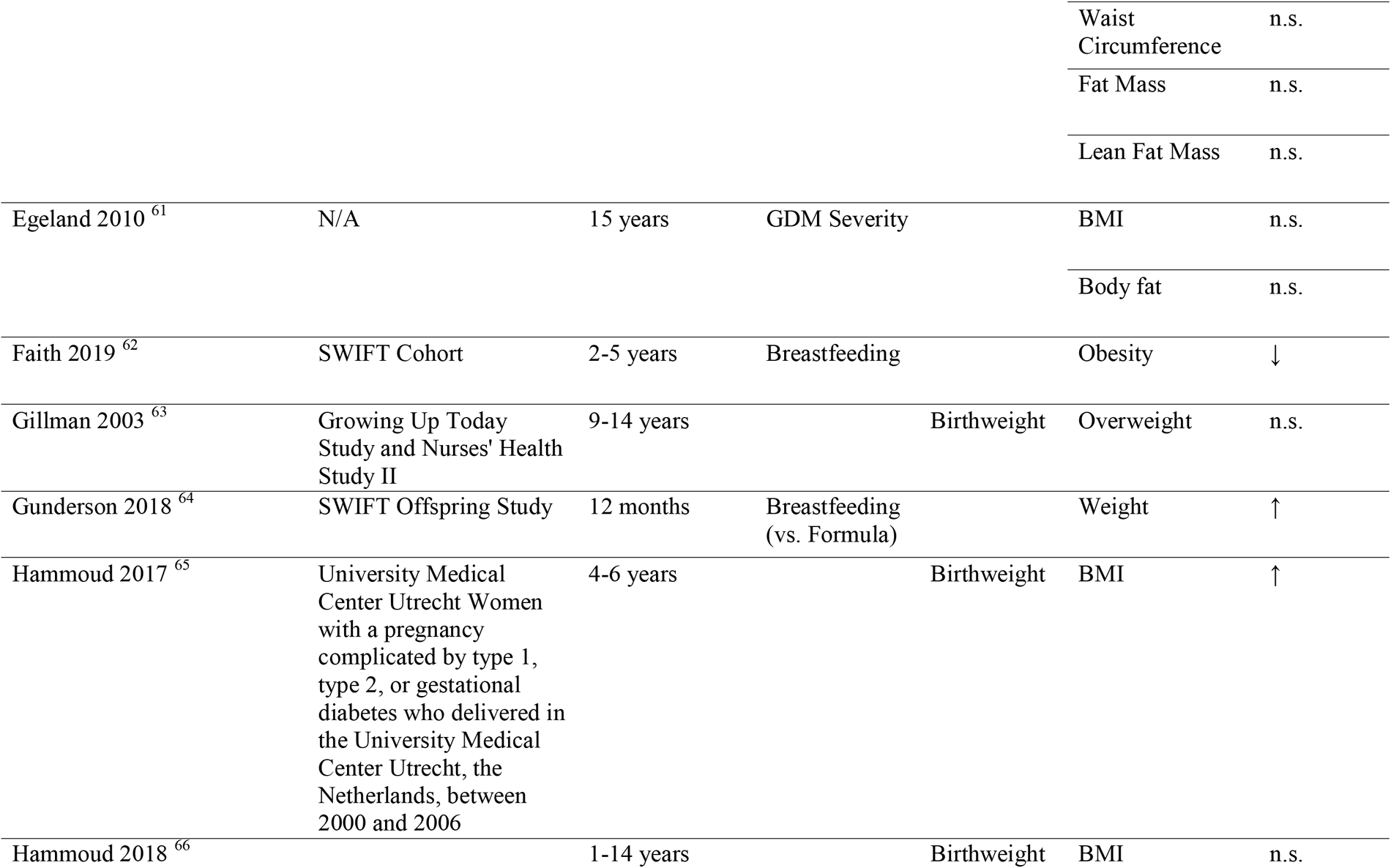
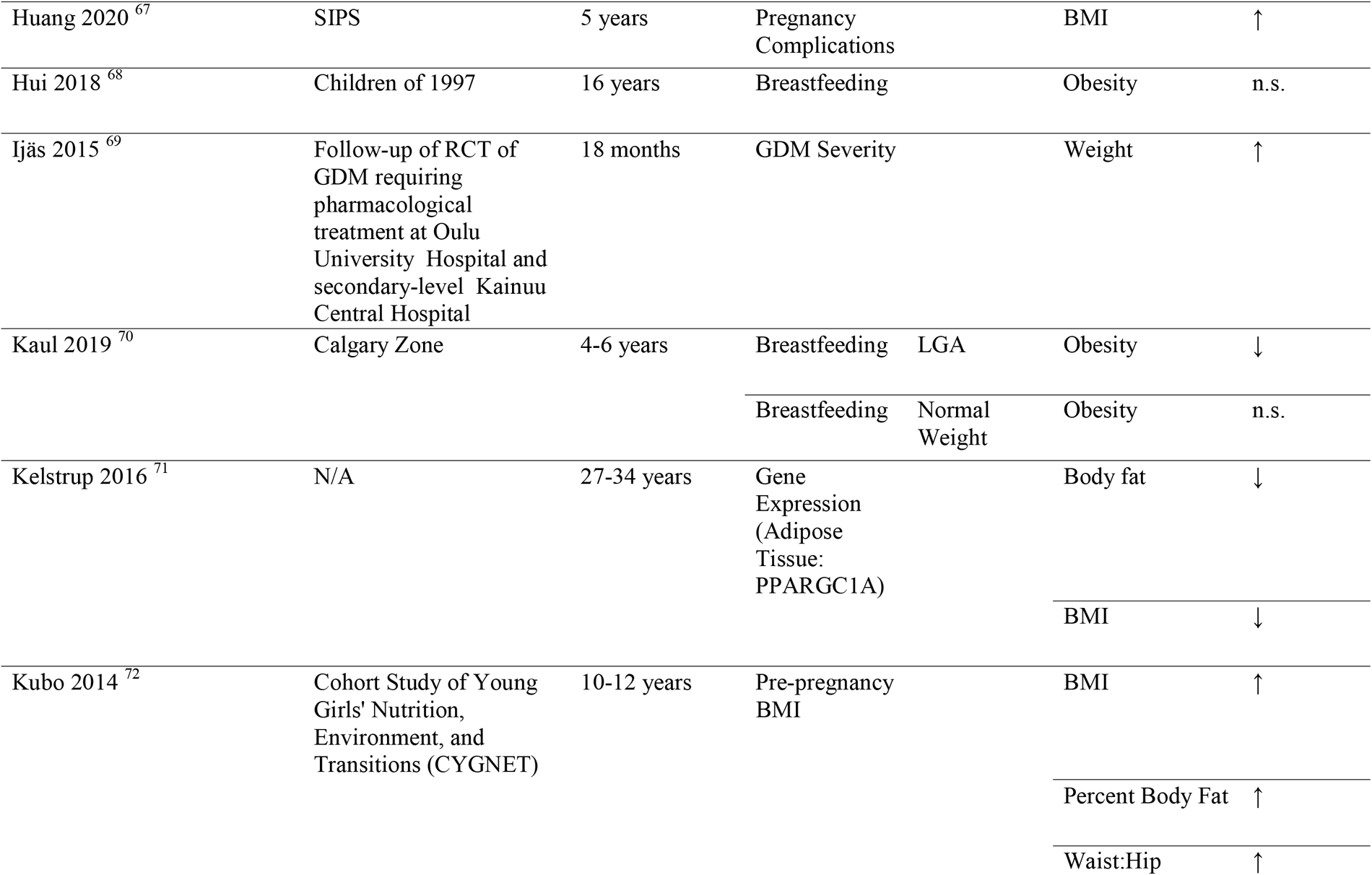
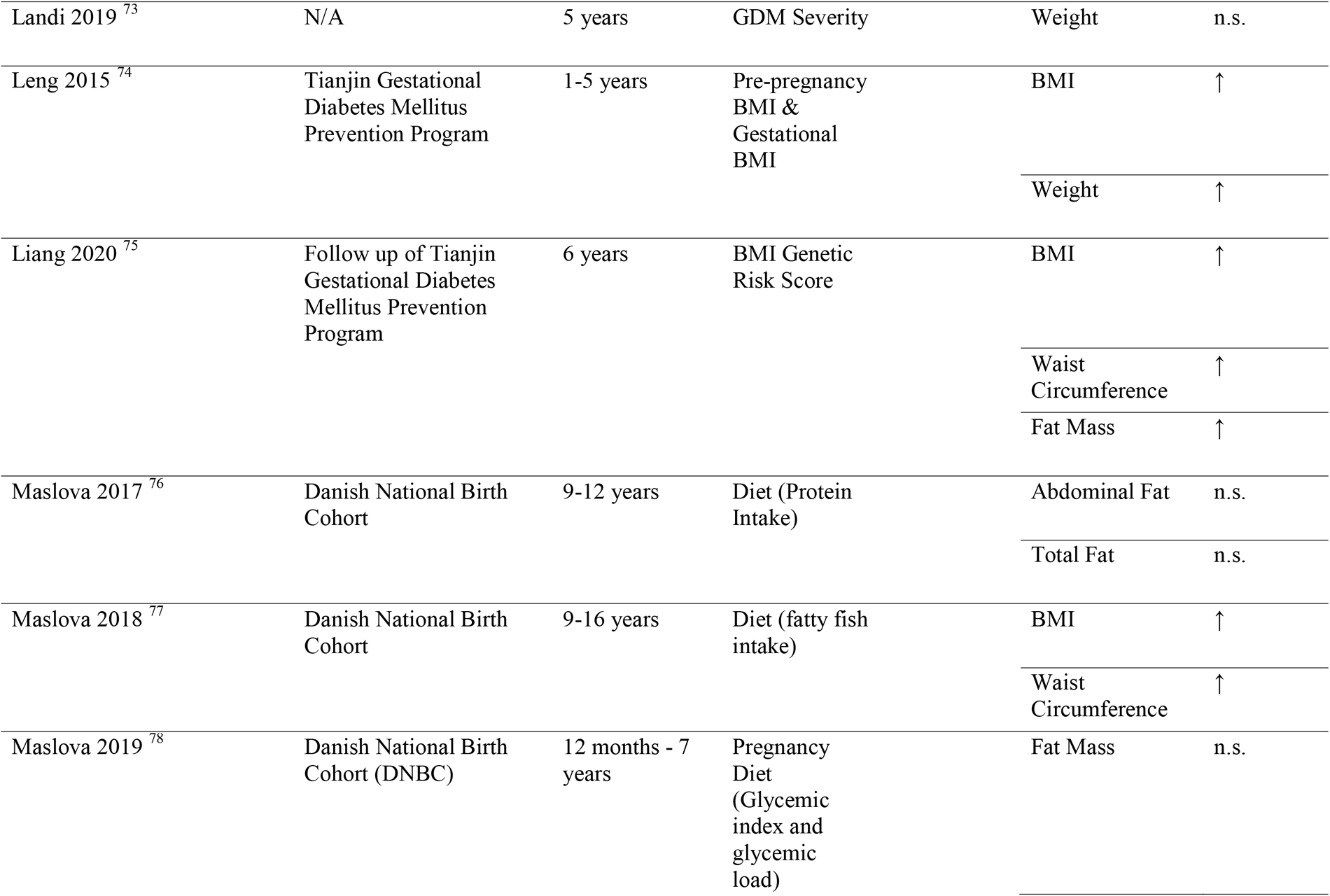
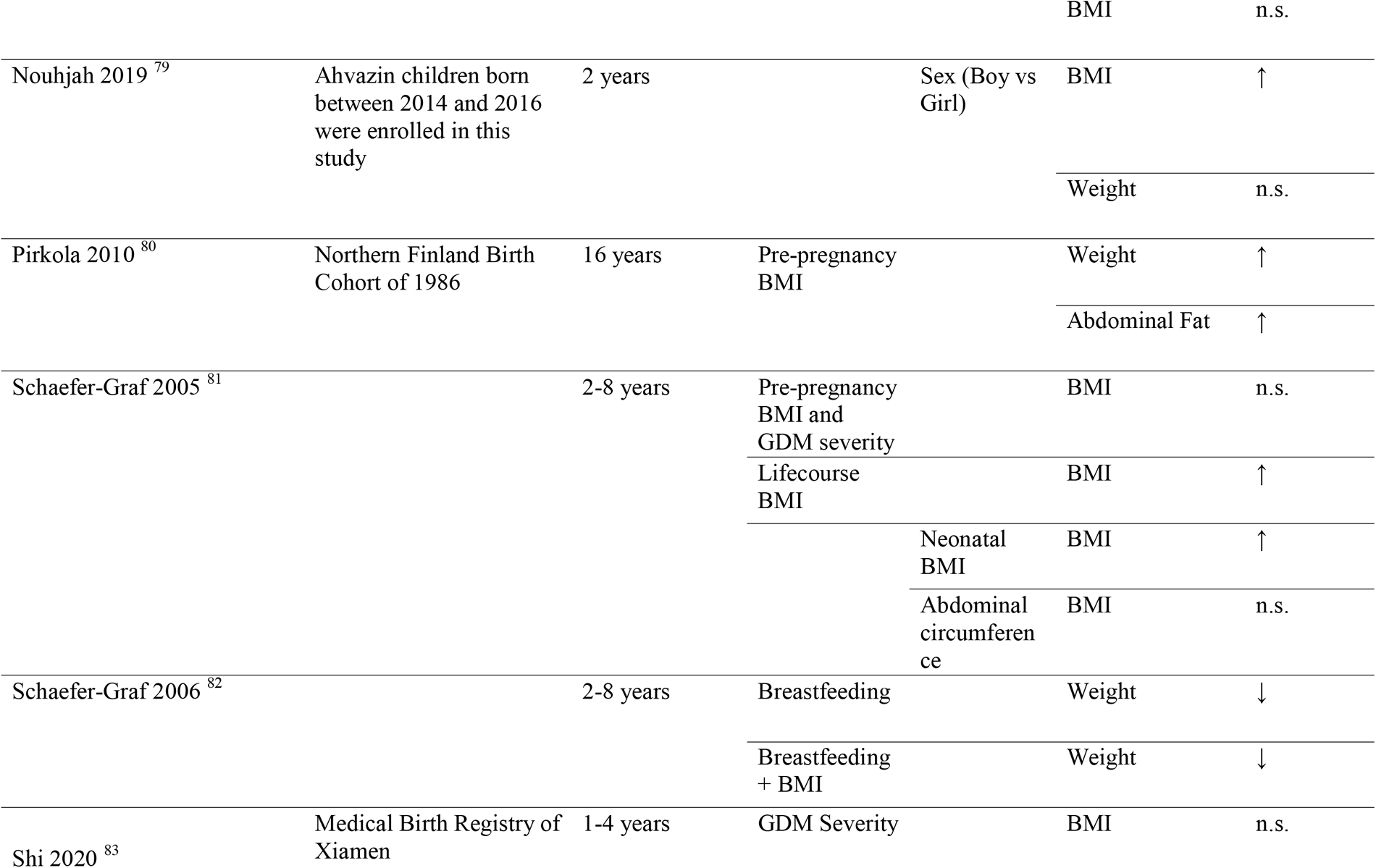
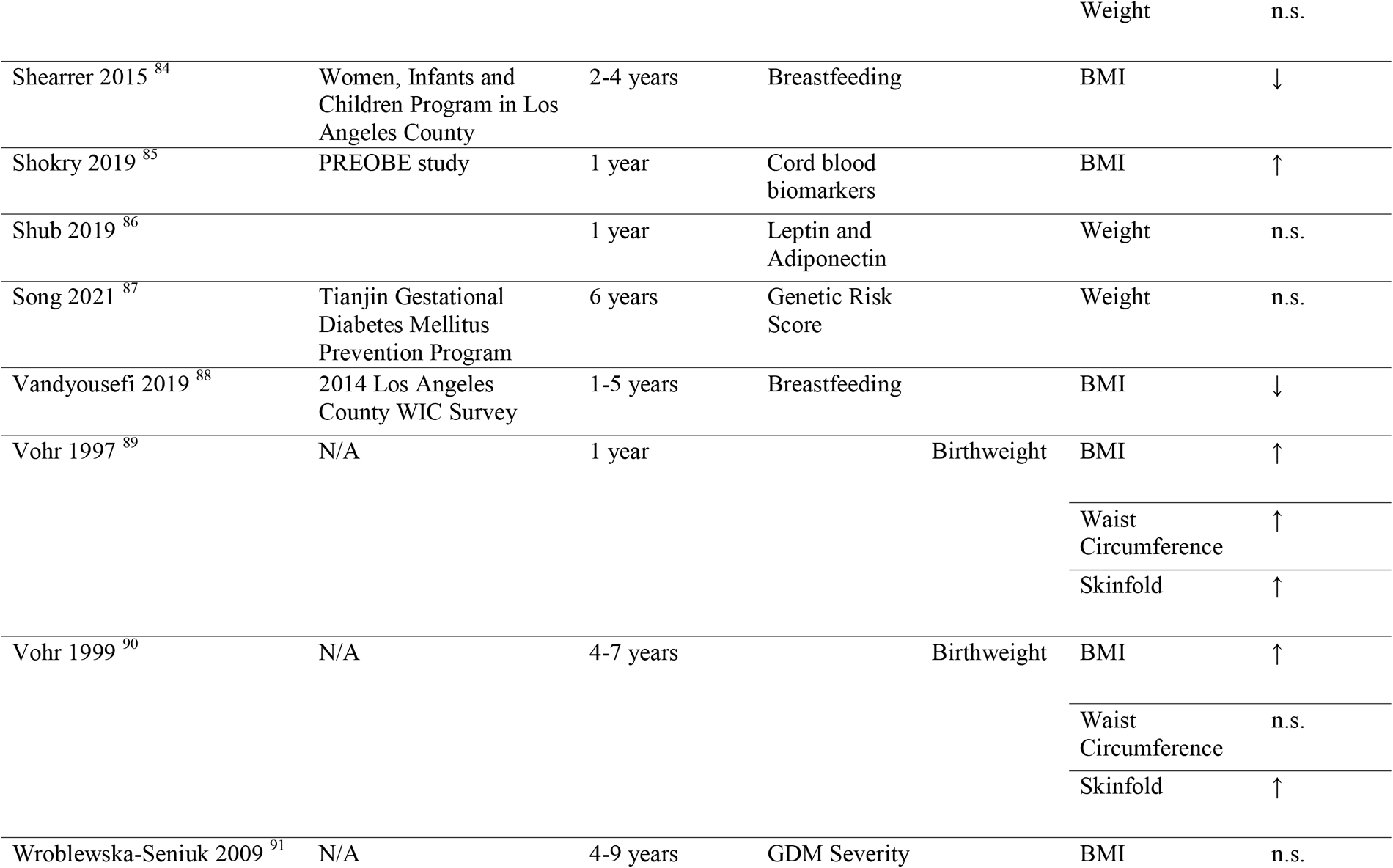
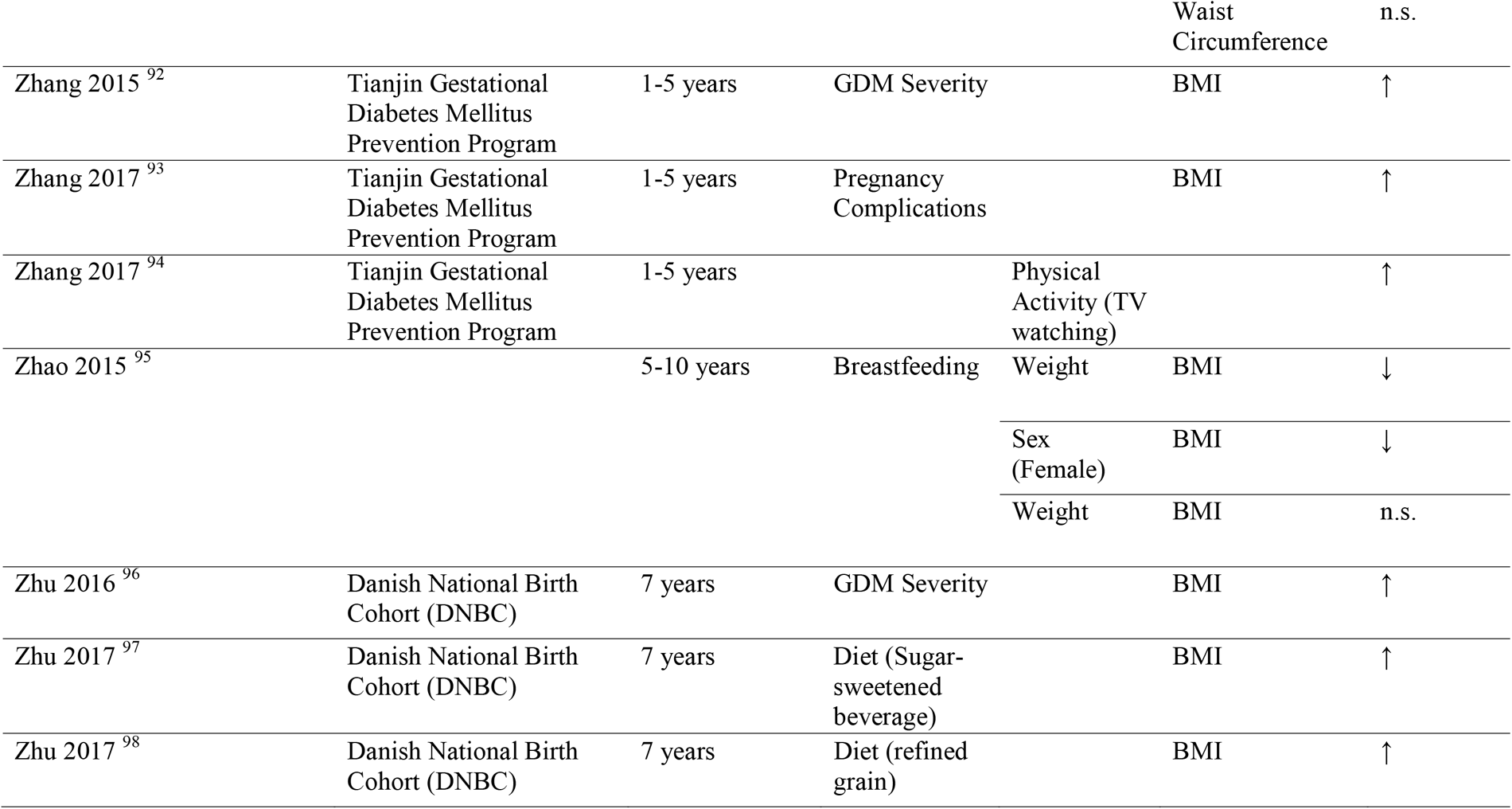
Observational studies evaluating prognostic factors in relation to anthropometric outcomes among offspring exposed to GDM *in utero* (*n*=43).

**TABLE 6.** Randomized control trials evaluating prognostic factors associated with anthropometric outcomes among offspring exposed to GDM in utero (n=4).

| Author Year | Trial Name | Intervention and/or Exposure | Offspring Outcome | Association |
| --- | --- | --- | --- | --- |
| Rowan 2011<sup>115</sup> | Metformin in Gestational diabetes (MiG): the offspring follow-up (TOFU) | Treatment (Metformin vs Insulin) | Skinfold | ↑ |
|  |  |  | Waist circumference | n.s. |
|  |  |  | Total fat | n.s. |
| Gillman 2010<sup>116</sup> | Australian Carbohydrate Intolerance Study in Pregnant Women (ACHOIS) | Treatment (dietary advice, blood glucose monitoring, and insulin therapy) vs. Routine Care | BMI | n.s. |
| Landon 2015<sup>110</sup> | Eunice Kennedy Shriver National Institute of Child Health and Human Development Maternal-Fetal Medicine Units (MFMU) Network randomized clinical trial (RCT) | Treatment (dietary advice, blood glucose monitoring, and insulin therapy) vs. Routine Care | BMI | n.s. |
|  |  |  | Waist circumference | n.s. |
| Malcolm 2006<sup>113</sup> | N/A | Tight vs minimal intervention | Weight | n.s. |

#### 3.2.2. Cardiometabolic profile

Only a few studies explored risk factors for adverse cardiometabolic outcomes in GDM offspring. These studies focused on blood pressure, lipids, and glycemic markers in the offspring separately or on metabolic syndrome like phenotypes, integrating several of these components and offspring anthropometric measures. Fourteen observational studies ^69–87,11^ (**Table 7**) and seven RCTs^82–88^ (**Table 8**) were identified. Four observational studies assessed associations of different maternal dietary components including macronutrients or specific micronutrients, and showed no consistent associations with offspring cardiometabolic outcomes. Several RCTs compared diet vs insulin treatment of GDM and showed no significant associations with the development of a metabolic syndrome like phenotype in the offspring. One RCT evaluating a lifestyle intervention including exercise and diet counselling for treatment of GDM compared to clinical care showed a higher risk of unfavorable metabolic outcomes in the offspring in the intervention group. One observational study showed that breastfeeding was associated with a lower risk of metabolic syndrome like phenotype in the offspring of GDM women, but one observational study showed no consistent effects. Birthweight was the most commonly-studied predictor of the offspring exposures. Two observational studies showed that a higher birthweight in offspring of GDM women was associated with a higher risk of metabolic syndrome like phenotype in offspring, whereas one study showed no association.

**TABLE 7.**
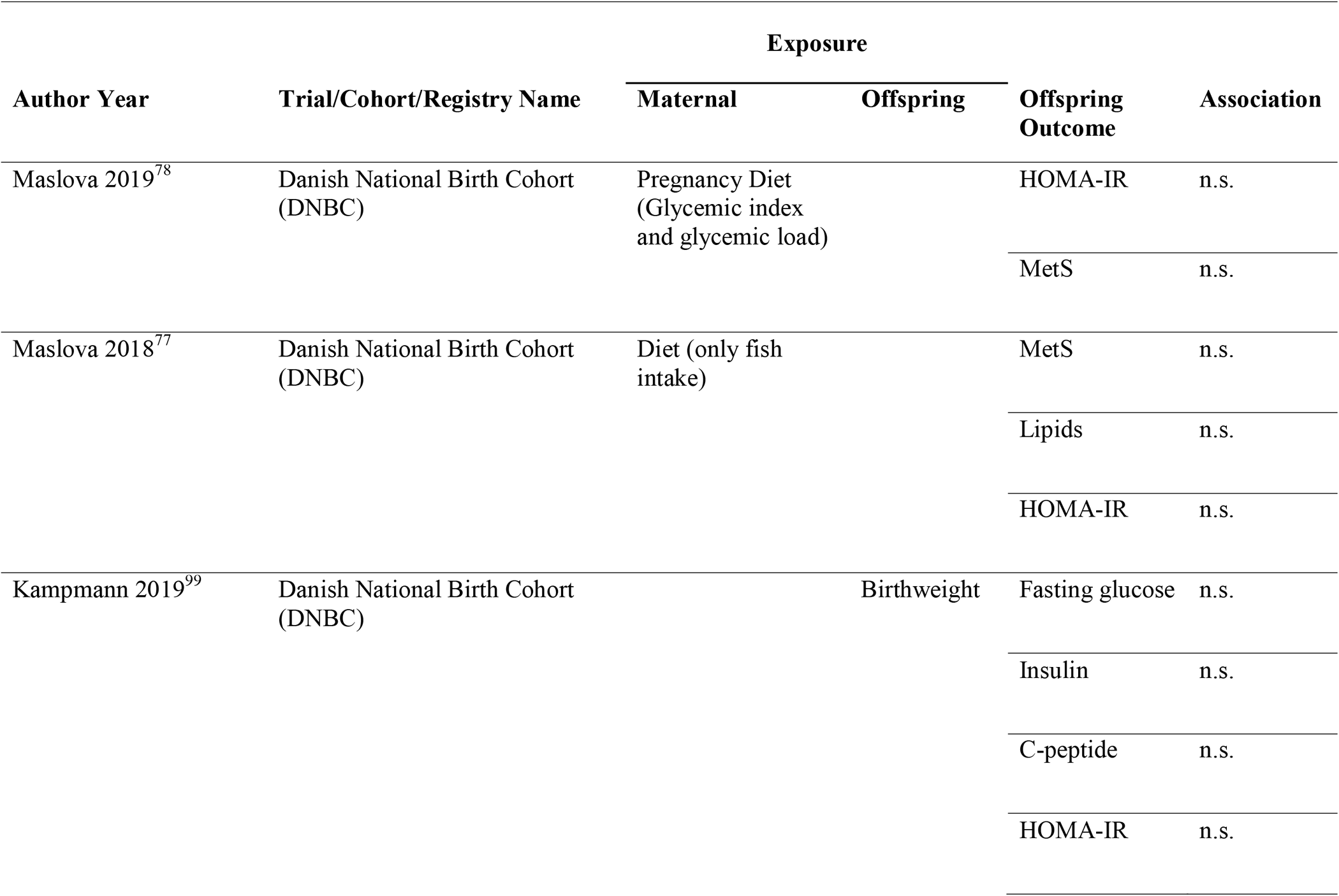
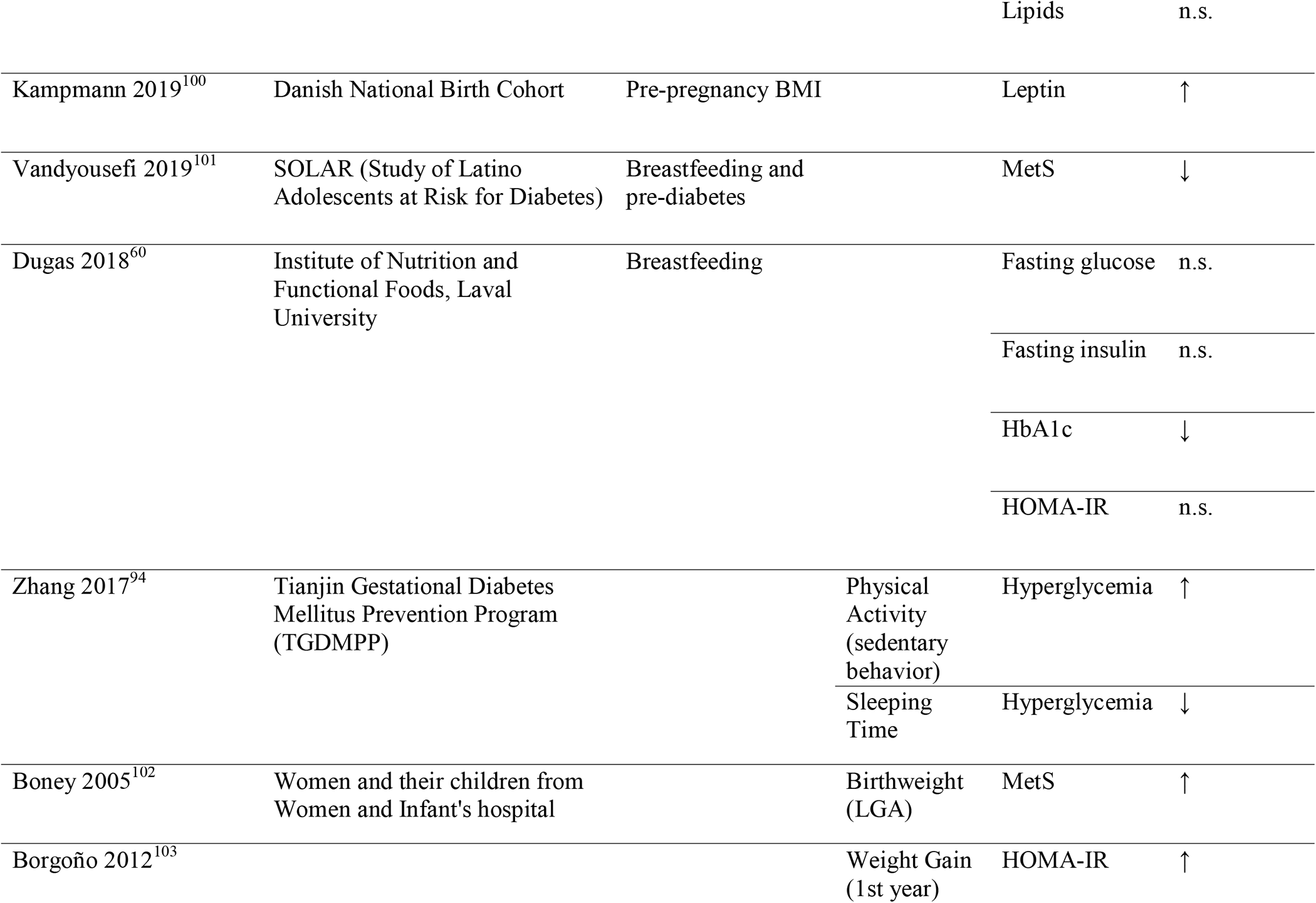
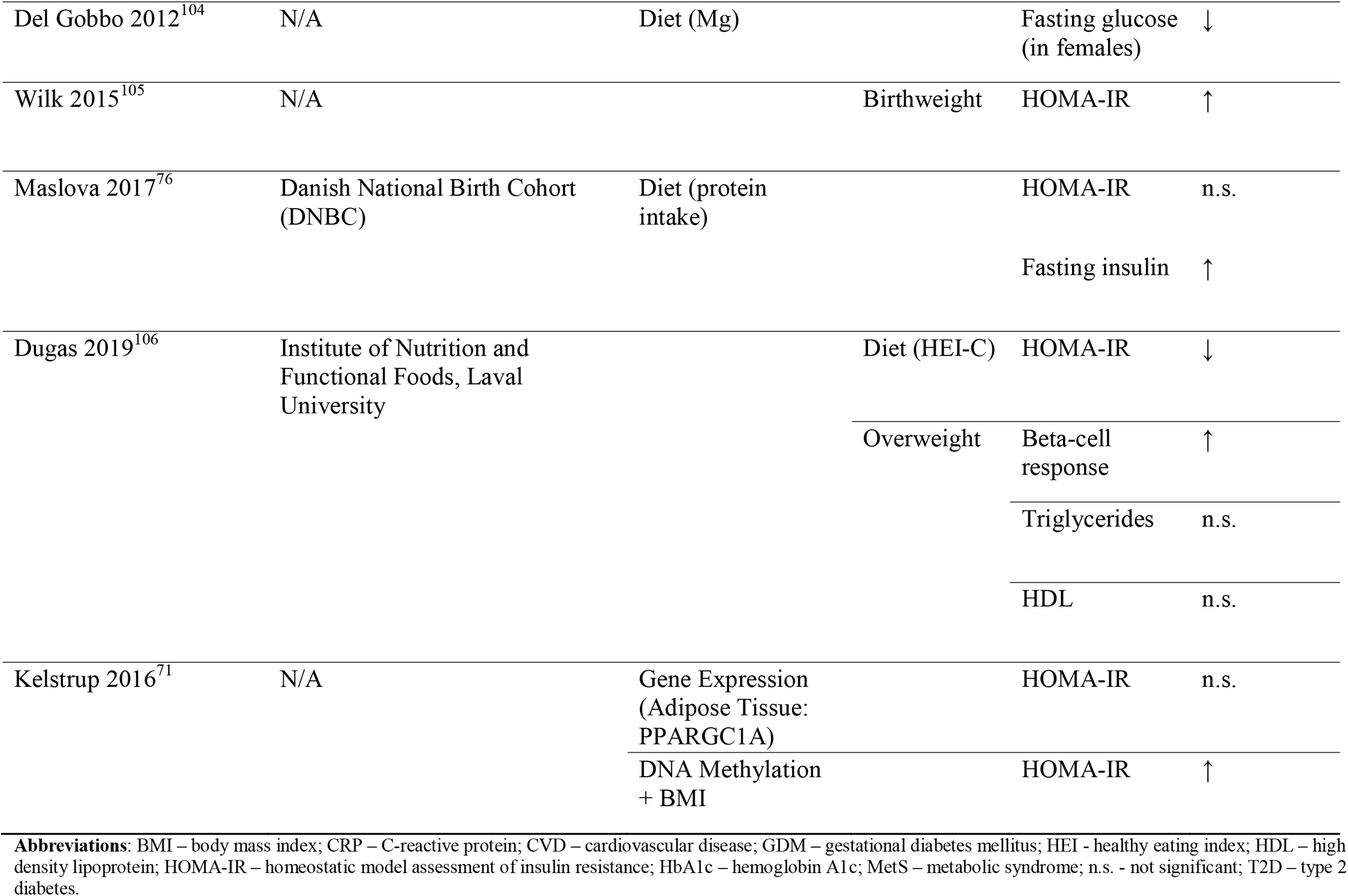
Observational studies evaluating prognostic factors in relation to cardiometabolic outcomes among offspring exposed to GDM *in utero* (*n*=14).

**TABLE 8.** Randomized control trials evaluating prognostic factors associated with cardiometabolic outcomes among offspring exposed to GDM in utero (n=7).

| Author Year | Trial Name | Intervention and/or Exposure | Offspring Outcome | Association |
| --- | --- | --- | --- | --- |
| Landon 2015<sup>110</sup> | Eunice Kennedy Shriver National Institute of Child Health and Human Development Maternal-Fetal Medicine Units (MFMU) Network randomized clinical trial (RCT) | Treatment (dietary intervention, self-monitory of glucose, and insulin therapy) vs. standard of care | Blood pressure | n.s. |
|  |  |  | Lipids | n.s. |
|  |  |  | HOMA-IR | n.s. |
|  |  | Offspring sex (male vs. female) | Fasting glucose | ↓ |
| Grotenfelt 2020<sup>111</sup> | Finnish Gestational Diabetes Prevention Study (RADIEL) | Lifestyle intervention (counseling of exercise and diet) | Metabolic risk profile | ↑ |
| Keely 2008<sup>112</sup> | RCT at the Ottawa Hospital (General and Civic campuses) and the Children's Hospital of Eastern Ontario, in Ottawa, Canada | Treatment (regular check-ups + dietary counseling) vs. Routine Care | Lipids | ↑ |
|  |  |  | HOMA-IR | ↑ |
|  |  |  | Glucose (fasting and 2 hour) | n.s. |
| Malcolm 2006<sup>113</sup> | RCT at the Ottawa Hospital (General and Civic campuses) and the Children's Hospital of Eastern Ontario, in Ottawa, | Treatment (regular check-ups and were given dietary counseling) vs. | Fasting glucose | n.s. |

|  | Canada | Routine Care |  |  |
| --- | --- | --- | --- | --- |
| Wroblewska-Seniuk 2009<sup>91</sup> | Children born in Clinical Hospital of Obstetrics and Gynecology in Poznan, Poland | Treatment (diet vs. insulin) | Glucose | n.s. |
|  |  |  | Insulin | n.s. |
|  |  |  | HOMA-IR | n.s. |
|  |  |  | HbA1c | n.s. |
| Egeland 2010<sup>61</sup> | N/A | Treatment (diet vs. insulin) | Insulin | ↓ |
|  |  |  | Glucose | ↓ |
|  |  |  | Fasting glucose | n.s. |
|  |  |  | Lipids | n.s. |
|  |  |  | HOMA-IR | n.s. |
| Battin 2015<sup>114</sup> | MiG Trial | Treatment (diet vs. insulin) | Blood pressure | n.s. |

**Abbreviations:** BMI – body mass index; CRP – C-reactive protein; CVD – cardiovascular disease; GDM – gestational diabetes mellitus; HOMA-IR – homeostatic model assessment of insulin resistance; HbA1c – hemoglobin A1c; : n.s. - not significant; T2D – type 2 diabetes.

#### 3.2.3. Quality of studies and certainty of evidence conducted in offspring populations

At the individual study level, we identified low quality of evidence for prognostic factors indicative of future adiposity and cardiometabolic risk among offspring exposed to GDM *in utero*. As with the maternal literature, most studies focusing on offspring outcomes were based on retrospective study designs leveraging registry data and observational cohort studies, both of which can be fraught with residual confounding and reverse causation bias, as well as structural biases like selection and attrition bias. Moreover, the literature of offspring outcomes remains relatively scant and with potentially inadequate durations of follow-up for manifestation of clinically-relevant cardiometabolic outcomes, though additional research is warranted. Furthermore, the certainty of evidence for maternal and offspring exposures with cardiometabolic outcomes were scored Level 4 based on the Diabetes Canada guidelines,^10^ based several factors including limited studies, small sample sizes, heterogeneity of study designs, and inadequate statistical methods.

## 4. DISCUSSION

### 4.1. Summary

This systematic review sought to identify prognostic risk factors during the perinatal period and across the lifecourse for maternal and offspring cardiovascular and metabolic outcomes among women and children affected by GDM pregnancies. We hypothesized that worse glycemic control at the time of GDM diagnosis (i.e., severity of GDM), older maternal age, belonging to racial/ethnic minority group; and unfavorable lifestyle behaviors would predict risk of full-blown type 2 diabetes (T2D) and cardiovascular disease (CVD) among women, and an unfavorable cardiometabolic profile among offspring.

The studies identified were primarily long-term retrospective and prospective studies. The level of evidence for prognostic risk factors of maternal T2D and CVD and for offspring cardiometabolic risk is low – i.e., Level 4 according to the Diabetes Canada 2018 Clinical Practice Guidelines for diabetes prognosis ^10^ – largely due to unmeasured confounding by lifestyle behaviors, the possibility of reverse causation bias due to pre-existing chronic conditions prior to or at the time of GDM diagnosis, and for offspring outcomes, the small body of literature on prognostic factors indicative of future adiposity and cardiometabolic risk. In the sections below, we synthesize general findings and comment further on quality of studies.

### 4.2. Maternal outcomes

#### 4.2.1. Cardiovascular disease

In line with a large literature demonstrating that women with a history of GDM are at higher risk CVD than their non-diabetic counterparts ^89, 90^, studies among women with a history of GDM indicated dose-response associations of GDM severity with these endpoints. Additionally, maternal pre-pregnancy or early pregnancy BMI was the most assessed prognostic factor linked to future CVD risk. Additionally, prenatal smoking, stillbirth, and gestational hypertension were each associated with risk for CVD in women with a history of GDM, whereas chronic hypertension and family history of CVD were not. Given the paucity of available research on CVD risk in women with a history of GDM, and the low certainty of evidence assessment, this is a research area ripe for investigation.

#### 4.2.2. Type 2 diabetes

Among women with GDM, higher BMI at any time in relation to the index pregnancy – i.e., pre-pregnancy, during the index pregnancy including gestational weight gain, and lifecourse measures of weight – predicted higher risk of T2D later in life. GDM severity, typically estimated by use of insulin or higher blood glucose values during the index pregnancy, was consistently associated with higher risk of developing T2D. While few studies assessed race and/or ethnicity as a prognostic risk factor, women of Asian or non-European descent with a history of GDM had higher risk of future T2D than White women ^24, 30, 50^. Breastfeeding duration and/or exclusivity was consistently associated with lower risk T2D risk following a GDM diagnosis during pregnancy, though follow-up often ended <2 years postpartum – a timeframe within which occult T2D incidence is relatively low. Longer duration follow-up is necessary to better evaluate the benefits of breastfeeding on T2D risk. Other lifestyle factors of interest included physical activity level during the perinatal and postpartum periods, as well as compliance with a healthy diet (e.g., adherence to a Mediterranean or DASH-like dietary pattern; the Healthy Eating Index score). However, observational findings with respect to these variables were mixed and RCTs investigating the effects of dietary interventions yielded mixed results ^62, 64^.

Several studies have examined biomarkers associated with increased risk for T2D following GDM pregnancies, including glycemic control at the time of GDM diagnosis, lipids ^28^, inflammatory biomarkers ^28^, and novel biomarkers lipidomics ^23^ and metabolomics ^47^ biomarkers. While these findings align with the literature in other pregnant ^91^ and non-pregnant populations ^92^, the low certainty of evidence from the studies and lack of replication/validation of findings prevent us from drawing firm conclusions regarding which factors may be the best predictors of future diabetes.

#### 4.2.3. Quality of maternal studies

We ranked the quality of evidence for prognostic factors indicative of risk of T2D or CVD in women as Level 4 (low) ^10^. Most empirical literature comes predominantly from large health care registries that boast large sample sizes and decades of follow-up. However, they carry high risk of bias in terms of identifying and interpretation specific prognostic characteristics as causal risk factors due to residual confounding due to maternal lifestyle, pre-existing chronic conditions, and other characteristics around time of pregnancy and GDM diagnoses. For example, maternal hypertension status in pregnancy as a causal risk factor for T2D or CVD could be explained in part by maternal BMI, diet quality, physical activity, smoking status, socioeconomic factors, and more. In contrast, there are notable large prospective cohorts, including the CARDIA study and the Nurses’ Health Study II, which collected detailed prospective information on the above-mentioned variables.

### 4.3. Offspring outcomes

#### 4.3.1. Anthropometry and body composition

The most common measure of offspring anthropometry was BMI between 2 and 10 years after birth. In alignment with maternal outcomes, GDM severity and higher maternal pre-pregnancy BMI were associated with higher offspring adiposity, as indicated by overweight/obesity status. Yet, interpretation of these findings should be tempered those of intervention studies showing that GDM treatment did not affect offspring anthropometrics ^82, 85, 93^. Similarly, RCTs testing the effects of pharmaceutical treatments (Metformin, insulin) and lifestyle interventions ^83^ to prevent GDM ^94, 95^. Other frequently studied perinatal predictors of offspring adiposity included birth size and breastfeeding duration/exclusivity. Generally, higher birthweight tended to be positively correlated with offspring BMI, though this association may be explained by the correlation between maternal and offspring BMI ^94^. Some observational studies showed a protective effect of breastfeeding on offspring childhood obesity risk, though this finding was not consistent and we did not identify any RCTs to support this finding. While a few studies reported a modifying effect of offspring sex on future body composition (e.g., ^96–98^), the direction of association was not consistent.

#### 4.3.2. Cardiometabolic profile

Most studies that assessed offspring cardiometabolic profile were observational and focused on prognostic factors that occurred during the perinatal/postpartum period, though we identified a few RCTs that targeted maternal glycemic control during pregnancy via pharmaceutical treatments and/or lifestyle alterations.

Among observational studies, the most common prognostic factor was GDM severity ^86, 99–101^ at times assessed in conjunction with maternal BMI ^100^. Although the magnitude of effects for GDM severity ranged across populations, greater severity was associated with worse glycemic control and an unfavorable cardiovascular profile in offspring. In addition to traditional biomarkers, one study identified hypothalamic activation as a determinant of BMI gain within the next year – a phenomenon that may mediate and/or modify the effect of GDM exposure *in utero* ^102^. As with the studies discussed above with respect to offspring anthropometry and body composition, RCTs to prevent GDM among high-risk women generally found minimal effects of the pharmaceutical and/or lifestyle interventions on offspring cardiometabolic profile, again suggesting that additional research is needed to better understand the pathophysiology of maternal GDM, to characterize relevant *in utero* programming pathways, and perhaps identify more specific and relevant offspring outcomes ^5^.

#### 4.3.3. Quality of offspring studies

As with the maternal studies, we categorized the literature on prognostic factors for offspring outcomes as being of low quality (Level 4). The inconsistent observational findings in conjunction with null results of RCTs targeting prevention of GDM among high-risk women indicate the existence of residual confounding for observational studies, and in the cases of the trials, the possibility that the interventions were developed with a suboptimal endpoint. Specifically, many prenatal interventions among women at high risk of GDM were developed with the goal of preventing macrosomia, which is a relatively crude indicator of developmental overnutrition and offspring adiposity. Future work is needed to gain a better understanding of in utero programming mechanisms that may link maternal GDM to offspring adiposity, as well as *de novo* interventions formulated to prevent neonatal adiposity assessed via gold standard methods such as air displacement plethysmography or dual X-ray absorptiometry ^5^.

## 5. STRENGTHS AND LIMITATIONS

### 5.1. Strengths and limitations of studies included in the systematic review

A key strength of many studies included in this systematic review is the prospective study design, which enhances temporal and causal inference regarding prognostic capacity of the maternal and offspring characteristics and behaviors assessed in studies herein. Additional strengths of some, but not all studies, include multi-ethnic study populations, which enhance generalizability of findings; large sample sizes, which improves capacity to detect biologically relevant associations; and use of gold standard assessments of the maternal and offspring outcomes of interest.

Limitations include the low-grade quality of studies included in this review (residual confounding, reverse causation bias, attrition and selection bias, inadequate duration of follow-up). Additionally, most studies were not designed to explore the long-term prognosis of GDM. Accordingly, many studies comprise *post hoc* analyses that were likely underpowered to detect smaller but biologically relevant effects of prognostic risk factors solely among mothers and/or offspring exposed to GDM. *Nota bene*, when screening studies, we noted that a general limitation of the literature on GDM prognostics in relation to offspring outcomes is assessment of the prognostic variable(s) contemporaneously with outcome assessment. These shortcomings resulted in high risk of bias and low quality of studies reviewed herein.

### 5.2. Strengths and limitations of systematic review approach and methodology

Strengths of the methodology for this systematic review include implementation of at least two independent reviews across all phases of the extraction and assessment process, with an additional review by a third independent reviewer to resolve conflicts; and adherence to well-established assessments of research quality and assessments of bias. Limitations include the exclusively qualitative synthesis of results – a necessity given the relatively small number of studies identified; and as with all systematic reviews, the potential for our conclusions to be impacted by publication bias.

## 6. FUTURE DIRECTIONS

Given the low quality of evidence identified in this systematic review, there is need for prospective cohort studies in diverse populations with granular data collection on prognostic risk factors as well as clinical and subclinical outcomes; high fidelity of follow-up across the lifecourse but, in particular, during sensitive windows of development during which there is greater developmental plasticity to respond to external cues ^103^; consideration of appropriate adjustment covariates depending on the specific prognostic risk factor of interest (e.g., there is discourse regarding whether maternal pre-pregnancy BMI should be included as a covariate in models where GDM severity is the prognostic risk factors of interest given that these two variables likely share overlapping *in utero* programming pathways ^117, 118^); and appropriate causal inference ^104^ and analytical approaches ^105^ to address structural biases that afflict observational study designs.

As interest in the application of precision prognostics to improve health for women and children affected by GDM pregnancies grows, there remains a need to establish a foundational knowledge base regarding traditional prognostic factors which, in turn, will enhance our ability to identify novel prognostic biomarkers that improve risk stratification for unfavorable health outcomes among mother-offspring dyads affected by GDM.

## Data Availability

N/A - this is a systematic review of published papers.

## ACKNOWLEDGEMENTS

The ADA/EASD Precision Diabetes Medicine Initiative, within which this work was conducted, has received the following support: The Covidence license was funded by Lund University (Sweden) for which technical support was provided by Maria Björklund and Krister Aronsson (Faculty of Medicine Library, Lund University, Sweden). Administrative support was provided by Lund University (Malmö, Sweden), University of Chicago (IL, USA), and the American Diabetes Association (Washington D.C., USA). The Novo Nordisk Foundation (Hellerup, Denmark) provided grant support for in-person writing group meetings (PI: L Phillipson, University of Chicago, IL). We also thank Marie-France Hivert for her valuable insights and feedback on this work.

## Abbreviations

BMI: body mass index
CRP: C-reactive protein
CVD: cardiovascular disease
GDM: gestational diabetes mellitus
MetS: metabolic syndrome
n.s.: not significant
SES: socioeconomic status
T2D: type 2 diabetes

