## Supplemental for "Predictors and risk factors of short-term and long-term outcomes among women with gestational diabetes mellitus (GDM) and their offspring: Moving toward precision prognosis?"

**Supplemental Table 1.** Search strategy.

|  |  | **Search Terms** |
| --- | --- | --- |
| maternal population | 1 | exp Diabetes, Gestational/ |
|  | 2 | gestational diabetes.mp. |
|  | 3 | GDM.mp. |
|  | 4 | diabetic pregnancy.mp. |
|  | 5 | (gestation$ adj4 diabet$).mp. [mp=title, abstract, original title, name of substance word, subject heading word, floating sub-heading word, keyword heading word, organism supplementary concept word, protocol supplementary concept word, rare disease supplementary concept word, unique identifier, synonyms] |
|  | 6 | (glucose adj4 (pregnan$ or gestation$ or prenatal$ or antenatal$ or pre-natal$ or ante-natal$ or maternal$)).mp. [mp=title, abstract, original title, name of substance word, subject heading word, floating sub-heading word, keyword heading word, organism supplementary concept word, protocol supplementary concept word, rare disease supplementary concept word, unique identifier, synonyms] |
|  | 7 | gestational diabet*.mp. |
|  | 8 | pregnancy induced diabetes.mp. |
|  | 9 | gestational diabetes [MeSH] |
| offspring population | 1 | exp Diabetes, Gestational/ |
|  | 2 | gestational diabetes.mp. |
|  | 3 | GDM.mp. |
|  | 4 | diabetic pregnancy.mp. |
|  | 5 | (gestation$ adj4 diabet$).mp. [mp=title, abstract, original title, name of substance word, subject heading word, floating sub-heading word, keyword heading word, organism supplementary concept word, protocol supplementary concept word, rare disease supplementary concept word, unique identifier, synonyms] |
|  | 6 | (glucose adj4 (pregnan$ or gestation$ or prenatal$ or antenatal$ or pre-natal$ or ante-natal$ or maternal$)).mp. [mp=title, abstract, original title, name of substance word, subject heading word, floating sub-heading word, keyword heading word, organism supplementary concept word, protocol supplementary concept word, rare disease supplementary concept word, unique identifier, synonyms] |
|  | 7 | gestational diabet*.mp. |
|  | 8 | pregnancy induced diabetes.mp. |
|  | 9 | 1 or 2 or 3 or 4 or 5 or 6 or 7 or 8 |
|  | 10 | offspring.mp. |
|  | 11 | exp Infant/ |
|  | 12 | infant.mp. |
|  | 13 | children.mp. |
|  | 14 | exp child/ |
|  | 15 | adolescent.mp. |
|  | 16 | exp adolescent/ |
|  | 17 | birth cohort.mp. |
|  | 18 | 10 or 11 or 12 or 13 or 14 or 15 or 16 or 17 |
|  | 19 | 9 and 18 |
|  | 20 | 9 or 19 |
| exposures/risk factors/predictors of prognosis | 21 | exp parity/ |
|  | 22 | parity.mp. |
|  | 23 | 21 or 22 |
|  | 24 | exp breast feeding/ |
|  | 25 | breastfeeding.mp. |
|  | 26 | 24 or 25 |
|  | 70 | exp Maternal Age/ |
|  | 71 | maternal age.mp. |
|  | 72 | 70 or 71 |
|  | 73 | exp gestational weight gain/ |
|  | 74 | gestational weight gain.mp. |
|  | 75 | 73 or 74 |
|  | 76 | exp fetal development/ |
|  | 77 | fetal growth.mp. |
|  | 78 | fetal development.mp. |
|  | 79 | 76 or 77 or 78 |
|  | 80 | exp pregnancy complications/ |
|  | 81 | pregnancy complications.mp. |
|  | 82 | 80 or 81 |
|  | 110 | exp Body Weight/ |
|  | 111 | body weight.mp. |
|  | 112 | (body mass index or bmi).mp. |
|  | 113 | body fat.mp. |
|  | 114 | adiposity.mp. |
|  | 115 | body size.mp. |
|  | 116 | waist circumference.mp. |
|  | 117 | waist-to-hip ratio.mp. |
|  | 118 | weight loss.mp. |
|  | 119 | weight gain.mp. |
|  | 120 | overweight.mp. |
|  | 121 | obesity.mp. |
|  | 122 | exp abdominal fat/ |
|  | 123 | exp Skinfold Thickness/ |
|  | 124 | exp Anthropometry/ |
|  | 125 | 110 or 111 or 112 or 113 or 114 or 115 or 116 or 117 or 118 or 119 or 120 or 121 or 122 or 123 or 124 |
|  | 27 | exp Blood Pressure/ |
|  | 28 | systolic blood pressure.mp. |
|  | 29 | SBP.mp. |
|  | 30 | diastolic blood pressure.mp. |
|  | 31 | DBP.mp. |
|  | 32 | exp hypertension/ |
|  | 33 | hypertension.mp. |
|  | 34 | high blood pressure.mp. |
|  | 35 | 27 or 28 or 29 or 30 or 31 or 32 or 33 or 34 |
|  | 36 | triacylglycerol*.mp. |
|  | 37 | triglyceride.mp. |
|  | 38 | VLDL.mp. |
|  | 39 | very low density lipoprotein.mp. |
|  | 40 | lipid*.mp. |
|  | 41 | exp lipids/ |
|  | 42 | exp cholesterol/ |
|  | 43 | cholesterol.mp. |
|  | 44 | lipoprotein.mp. |
|  | 45 | exp lipoproteins/ |
|  | 46 | (hdl or high density lipoprotein).mp. |
|  | 47 | (ldl or low density lipoprotein).mp. |
|  | 48 | exp hyperlipidemias/ |
|  | 49 | apolipoprotein*.mp. |
|  | 50 | non-hdl.mp. |
|  | 51 | lipidemia*.mp. |
|  | 52 | lipemia*.mp. |
|  | 53 | Lipemic.mp. |
|  | 54 | 36 or 37 or 38 or 39 or 40 or 41 or 42 or 43 or 44 or 45 or 46 or 47 or 48 or 49 or 50 or 51 or 52 or 53 |
|  | 55 | exp leptin/ |
|  | 56 | leptin.mp. |
|  | 57 | 55 or 56 |
|  | 58 | CRP.mp. |
|  | 59 | high-sensitivity CRP.mp. |
|  | 60 | c-reactive protein.mp. |
|  | 61 | hs-CRP.mp. |
|  | 62 | 58 or 59 or 60 or 61 |
|  | 63 | cpeptide.mp. |
|  | 64 | c-peptide.mp. |
|  | 65 | exp C-Peptide/ |
|  | 66 | 63 or 64 or 65 |
|  | 67 | exp adiponectin/ |
|  | 68 | adiponectin.mp. |
|  | 69 | 67 or 68 |
|  | 83 | glyc*m*.mp. |
|  | 84 | Hemoglobin A, Glycosylated/ |
|  | 85 | glyc*mia.mp. |
|  | 86 | insulin*.mp. |
|  | 87 | gly* albumin.mp. |
|  | 88 | OGTT.mp. |
|  | 89 | hba1c.mp. |
|  | 90 | HOMA*.mp. |
|  | 91 | fructosamine*.mp. |
|  | 92 | Insulin/ |
|  | 93 | exp Glucose/ |
|  | 94 | exp Glucose Tolerance Test/ |
|  | 95 | exp Hyperinsulinism/ |
|  | 96 | 83 or 84 or 85 or 86 or 87 or 88 or 89 or 90 or 91 or 92 or 93 or 94 or 95 |
|  | 97 | exp genomics/ |
|  | 98 | exp metabolomics/ |
|  | 99 | metabolomic*.mp. |
|  | 100 | metabolite.mp. |
|  | 101 | exp metabolome/ |
|  | 102 | metabolome.mp. |
|  | 103 | exp proteomics/ |
|  | 104 | proteomic*.mp. |
|  | 105 | genomic*.mp. |
|  | 106 | exp Epigenesis, Genetic/ |
|  | 107 | epigenetic*.mp. |
|  | 108 | epigenesis.mp. |
|  | 109 | 97 or 98 or 99 or 100 or 101 or 102 or 103 or 104 or 105 or 106 or 107 or 108 |
|  | 126 | family history.mp. |
|  | 127 | parental history.mp. |
|  | 128 | 126 or 127 |
|  | 129 | diabetes.mp. |
|  | 130 | diabetes mellitus.mp. |
|  | 131 | exp diabetes mellitus, type 2/ |
|  | 132 | T2D*.mp. |
|  | 133 | type 2 diabetes.mp. |
|  | 134 | 129 or 130 or 131 or 132 or 133 |
|  | 135 | 128 and 134 |
|  | 136 | exp diet/ |
|  | 137 | nutrition.mp. |
|  | 138 | diet*.mp. |
|  | 139 | exp dietary supplements/ |
|  | 140 | exp diet, carbohydrate-restricted/ |
|  | 141 | exp diet, fat-restricted/ |
|  | 142 | exp caloric restriction/ |
|  | 143 | food.mp. |
|  | 144 | diet therapy.mp. |
|  | 145 | diet* intervention*.mp. |
|  | 146 | macronutrient.mp. |
|  | 147 | supplement*.mp. |
|  | 148 | protein.mp. |
|  | 149 | meat.mp. |
|  | 150 | food*.mp. |
|  | 151 | beverage*.mp. |
|  | 152 | meal*.mp. |
|  | 153 | 136 or 137 or 138 or 139 or 140 or 141 or 142 or 143 or 144 or 145 or 146 or 147 or 148 or 149 or 150 or 151 or 152 |
|  | 154 | exp exercise/ |
|  | 155 | physical activity.mp. |
|  | 156 | exp physical fitness/ |
|  | 157 | physical fitness.mp. |
|  | 158 | 154 or 155 or 156 or 157 |
|  | 159 | exp "social determinants of health"/ |
|  | 160 | "social determinants of health".mp. |
|  | 161 | exp socioeconomic factors/ |
|  | 162 | exp health status disparities/ |
|  | 163 | exp Educational Status/ |
|  | 164 | exp Income/ |
|  | 165 | (socio-economic* or socioeconomic* or social determinant* or insurance status or standard* of living or lower income or under-insured or social class* or resource poor or social condition* or disadvantage* or social status or household income or family income or income level*).mp. |
|  | 166 | educational attainment.mp. |
|  | 167 | 159 or 160 or 161 or 162 or 163 or 164 or 165 or 166 |
| maternal outcomes | 168 | exp cardiovascular diseases/ |
|  | 169 | CVD.mp. |
|  | 170 | exp myocardial infarction/ |
|  | 171 | myocardial infarction.mp. |
|  | 172 | exp Stroke/ |
|  | 173 | stroke.mp. |
|  | 174 | heart failure.mp. |
|  | 175 | exp heart failure/ |
|  | 176 | coronary disease.mp. |
|  | 177 | exp coronary disease/ |
|  | 178 | myocardial isch*.mp. |
|  | 179 | ckd.mp. |
|  | 180 | exp Renal Insufficiency, Chronic/ |
|  | 181 | exp myocardial ischemia/ |
|  | 182 | chronic kidney disease.mp. |
|  | 183 | 168 or 169 or 170 or 171 or 172 or 173 or 174 or 175 or 176 or 177 or 178 or 179 or 180 or 181 or 182 |
| offspring outcomes | 110 | exp Body Weight/ |
|  | 111 | body weight.mp. |
|  | 112 | (body mass index or bmi).mp. |
|  | 113 | body fat.mp. |
|  | 114 | adiposity.mp. |
|  | 115 | body size.mp. |
|  | 116 | waist circumference.mp. |
|  | 117 | waist-to-hip ratio.mp. |
|  | 118 | weight loss.mp. |
|  | 119 | weight gain.mp. |
|  | 120 | overweight.mp. |
|  | 121 | obesity.mp. |
|  | 122 | exp abdominal fat/ |
|  | 123 | exp Skinfold Thickness/ |
|  | 124 | exp Anthropometry/ |
|  | 125 | 110 or 111 or 112 or 113 or 114 or 115 or 116 or 117 or 118 or 119 or 120 or 121 or 122 or 123 or 124 |
|  | 27 | exp Blood Pressure/ |
|  | 28 | systolic blood pressure.mp. |
|  | 29 | SBP.mp. |
|  | 30 | diastolic blood pressure.mp. |
|  | 31 | DBP.mp. |
|  | 32 | exp hypertension/ |
|  | 33 | hypertension.mp. |
|  | 34 | high blood pressure.mp. |
|  | 35 | 27 or 28 or 29 or 30 or 31 or 32 or 33 or 34 |
|  | 36 | triacylglycerol*.mp. |
|  | 37 | triglyceride.mp. |
|  | 38 | VLDL.mp. |
|  | 39 | very low density lipoprotein.mp. |
|  | 40 | lipid*.mp. |
|  | 41 | exp lipids/ |
|  | 42 | exp cholesterol/ |
|  | 43 | cholesterol.mp. |
|  | 44 | lipoprotein.mp. |
|  | 45 | exp lipoproteins/ |
|  | 46 | (hdl or high density lipoprotein).mp. |
|  | 47 | (ldl or low density lipoprotein).mp. |
|  | 48 | exp hyperlipidemias/ |
|  | 49 | apolipoprotein*.mp. |
|  | 50 | non-hdl.mp. |
|  | 51 | lipidemia*.mp. |
|  | 52 | lipemia*.mp. |
|  | 53 | Lipemic.mp. |
|  | 54 | 36 or 37 or 38 or 39 or 40 or 41 or 42 or 43 or 44 or 45 or 46 or 47 or 48 or 49 or 50 or 51 or 52 or 53 |
|  | 55 | exp leptin/ |
|  | 56 | leptin.mp. |
|  | 57 | 55 or 56 |
|  | 58 | CRP.mp. |
|  | 59 | high-sensitivity CRP.mp. |
|  | 60 | c-reactive protein.mp. |
|  | 61 | hs-CRP.mp. |
|  | 62 | 58 or 59 or 60 or 61 |
|  | 63 | cpeptide.mp. |
|  | 64 | c-peptide.mp. |
|  | 65 | exp C-Peptide/ |
|  | 66 | 63 or 64 or 65 |
|  | 67 | exp adiponectin/ |
|  | 68 | adiponectin.mp. |
|  | 69 | 67 or 68 |
|  | 83 | glyc*m*.mp. |
|  | 84 | Hemoglobin A, Glycosylated/ |
|  | 85 | glyc*mia.mp. |
|  | 86 | insulin*.mp. |
|  | 87 | gly* albumin.mp. |
|  | 88 | OGTT.mp. |
|  | 89 | hba1c.mp. |
|  | 90 | HOMA*.mp. |
|  | 91 | fructosamine*.mp. |
|  | 92 | Insulin/ |
|  | 93 | exp Glucose/ |
|  | 94 | exp Glucose Tolerance Test/ |
|  | 95 | exp Hyperinsulinism/ |
|  | 96 | 83 or 84 or 85 or 86 or 87 or 88 or 89 or 90 or 91 or 92 or 93 or 94 or 95 |
|  | 184 | fatty liver.mp. |
|  | 185 | exp non-alcoholic fatty liver disease/ |
|  | 186 | NAFLD.mp. |
|  | 187 | exp fatty liver/ |
|  | 188 | 184 or 185 or 186 or 187 |
|  | 189 | metabolic syndrome.mp. |
|  | 190 | syndrome x.mp. |
|  | 191 | cardio-metabolic syndrome.mp. |
|  | 192 | MetS.mp. |
|  | 193 | exp metabolic syndrome/ |
|  | 194 | 189 or 190 or 191 or 192 or 193 |
|  | 168 | exp cardiovascular diseases/ |
|  | 169 | CVD.mp. |
|  | 170 | exp myocardial infarction/ |
|  | 171 | myocardial infarction.mp. |
|  | 172 | exp Stroke/ |
|  | 173 | stroke.mp. |
|  | 174 | heart failure.mp. |
|  | 175 | exp heart failure/ |
|  | 176 | coronary disease.mp. |
|  | 177 | exp coronary disease/ |
|  | 178 | myocardial isch*.mp. |
|  | 179 | ckd.mp. |
|  | 180 | exp Renal Insufficiency, Chronic/ |
|  | 181 | exp myocardial ischemia/ |
|  | 182 | chronic kidney disease.mp. |
|  | 183 | 168 or 169 or 170 or 171 or 172 or 173 or 174 or 175 or 176 or 177 or 178 or 179 or 180 or 181 or 182 |
|  | 195 | exp Cohort Studies/ |
|  | 196 | cohort.mp. |
|  | 197 | cohort$.tw. |
|  | 198 | controlled clinical trial.pt. |
|  | 199 | epidemiologic methods/ |
|  | 200 | exp Clinical Trial/ |
|  | 201 | exp medical records/ |
|  | 202 | 195 or 196 or 197 or 198 or 199 or 200 or 201 |
|  | 203 | 9 or 19 |
|  | 204 | 23 or 26 or 35 or 54 or 57 or 62 or 66 or 69 or 72 or 75 or 79 or 82 or 96 or 109 or 125 or 135 or 153 or 158 or 167 |
|  | 205 | 54 or 57 or 62 or 66 or 69 or 96 or 125 or 134 or 183 or 188 or 194 |
|  | 206 | 202 and 203 and 204 and 205 |
|  | 207 | limit 206 to (english language and humans and yr="1990 -Current") |

**Supplemental Table 2.** Diabetes Canada 2018 Clinical Practice Guidelines

| Studies of treatment and prevention | |
| --- | --- |
| Level 1A | Systematic overview or meta-analysis of high-quality RCTs   a. Comprehensive search for evidence a. Authors avoided bias in selecting articles for inclusion a. Authors assessed each article for validity a. Reports clear conclusions that are supported by the data and appropriate analyses   OR Appropriately designed RCT with adequate power to answer the question posed by the investigators   a. Patients were randomly allocated to treatment groups a. Follow up at least 80% complete a. Patients and investigators were blinded to the treatment ^∗^ a. Patients were analyzed in the treatment groups to which they were assigned a. The sample size was large enough to detect the outcome of interest |
| Level 1B | Non-randomized clinical trial or cohort study with indisputable results |
| Level 2 | RCT or systematic overview that does not meet Level 1 criteria |
| Level 3 | Non-randomized clinical trial or cohort study; systematic overview or meta-analysis of level 3 studies |
| Level 4 | Other |
